# Partner-inflicted brain injury may cognitively age survivors by over a decade

**DOI:** 10.64898/2026.09.24.26363984

**Authors:** Craig W. McFarland, Eve M. Valera

**Affiliations:** Department of Experimental Psychology, University of Oxford; Oxford OX2 6GG, UK; Stanford Interdepartmental Neurosciences Program, Stanford University School of Medicine; Stanford, CA 94305, USA; Department of Psychiatry, Mass General Brigham; Boston, MA 02129, USA; Department of Psychiatry, Harvard Medical School; Boston, MA 02115, USA

## Abstract

**Introduction:** More than a third of women in the United States are estimated to experience abuse by an intimate partner, and blows to the head and strangulation are common in such instances. Yet the cognitive consequences of partner-inflicted brain injuries remain poorly characterized relative to brain injuries from other mechanisms, such as athletics and military service, and study sample sizes have tended to be small. The degree to which these injuries accumulate into measurable cognitive harm, whether the harm of such injuries can be quantified in a meaningful way, and how that harm compares with better-recognized causes of repetitive head injury, has not been established.

**Rationale:** We administered ten cognitive tests, scored across sixteen domains, to 207 women with lifetime exposure to physical intimate partner violence, of whom 67% (*n* = 139) reported at least one partner-inflicted brain injury. Since standardized scores are difficult to interpret clinically, we also expressed performance as cognitive age, the age at which the typical person in the general population performs equivalently. Finally, we compared women with brain injuries against women with no brain injury history from the same cohort, as this isolates injury from exposure to violence itself, since both groups experienced physical partner violence.

**Results:** Women who had sustained any partner-inflicted brain injury performed below the general population mean on 11 of the 16 cognitive abilities measured, from reasoning (*d* = 0.32) to reaction time (*d* = 0.80), all *P* ≤ 0.011. Women reporting ten or more injuries demonstrated an overall cognitive function worse than nearly 80% of the general population (*d* = 0.82, *P* < 0.0001). Translating these deficits to cognitive age, women with ten or more brain injuries performed at a cognitive age more than two decades older than the general population on eight domains, the largest gaps being for simple attention (28.1 years, 95% CI [14.9, 41.4]; *d* = 0.78, *P* = 0.0002) and reaction time (27.8 years, 95% CI [18.1, 37.4]; *d* = 1.08, *P* < 0.0001). On psychomotor speed they also performed over a decade older than women exposed to physical intimate partner violence who had never sustained a brain injury (11.3 years, *P* = 0.033). After adjustment for eleven covariates, cognitive ability declined as injury count rose in six domains: psychomotor speed, reaction time, processing speed, motor speed, global cognition and verbal memory (partial *r* = −0.14 to −0.22, all *P* < 0.05), and 14 of the 16 coefficients were negative (two-sided sign test *P* = 0.004). We excluded scores flagged as invalid by the cognitive assessment per convention, yet observed that these flags fell disproportionately on the most impaired women, who scored 11 to 13 points lower (*d* = 0.60 to 0.84, all *P* < 0.001) on every domain the assessment did accept as valid.

**Conclusion:** Women experiencing partner-inflicted brain injuries demonstrated cognitive aging of over 20 years, and yet, the true cognitive harms of such injury are likely more severe than those reported here. These results are particularly grave because partner-inflicted injuries may compromise the very cognitive capacities women must rely on to leave abusive relationships, care for their children, sustain employment, navigate legal proceedings, and remain safe post-violence. Taken together, these findings underscore partner-inflicted brain injury as a major, under-recognized source of cognitive harm, potentially affecting more women than brain injury in professional football or the military while lacking comparable clinical recognition and care.

**Summary Abstract:** More than a third of women in the United States are estimated to experience abuse from an intimate partner, in which blows to the head, strangulation, and therefore brain injuries are common. Among 207 survivors of such physical violence, two-thirds reported a partner-inflicted brain injury. These women performed substantially worse than the general population across most cognitive measures, including memory and reaction time. Cognitive performance declined as injury count increased. Translated into cognitive age, survivors with the most partner-inflicted brain injuries performed over two decades older than the general population, and a decade older than survivors with no history of brain injury. Importantly, the cognitive assessment discards scores it judges unreliable, yet these came primarily from the most impaired women, suggesting that the true harm is likely greater than reported here. Such cognitive deficits may limit these women’s capacity to live to their potential, including keeping safe from abuse. These findings underscore partner-inflicted brain injury as a major, under-recognized source of cognitive harm, potentially affecting more women than brain injury in professional football or the military while lacking comparable clinical recognition and care.

## Introduction

More than a third of women in the United States are estimated to experience abuse from an intimate partner [1]. The threat of abuse intensifies in response to current events beyond their control, with rates reported rising during war [2,3], recessions [4,5], pandemic restrictions [6,7], floods [8,9], hurricanes [10–12], heatwaves [13,14], the Super Bowl [15], the World Cup [16], when the weekend comes [15,17], and, among other holidays, on Christmas [17].

Intimate partner violence (IPV) is self-evidently concerning, but a particularly troubling consequence is brain injury (BI). During abuse, survivors experience blows, kicks, falls, strangulations, and high-force impacts against walls or floors that can produce BIs [18–21]. The harms of BI compound, particularly as this “invisible trauma” can occur repeatedly without diagnosis, time to recover, or treatment for survivors of intimate partner violence [18,19]. The cognitive detriments of BI are frequently mistaken for psychological distress caused by the abuse, leaving the underlying BI unrecognized [18,20].

Heightened risks of stroke [22], seizures [23], and death [24] notwithstanding, BI impairs cognitive functions, often to debilitating extents [19,25–28]. Accordingly, billions of dollars have been invested in BI research among athletes and military personnel, informing legislation, clinical guidance, and safety initiatives [29–32]. Intimate partner violence has no comparable counterpart, although partner-inflicted BI is common, frequently repetitive, and may affect more people than BI in these traditionally studied populations [19,20,33]. Characterizing the breadth and severity of the cognitive harms from partner-inflicted BI is therefore crucial to understanding the impact of IPV, and consequently informing effective clinical care and relevant legislation.

Here we studied 207 women who had experienced physical violence from an intimate partner, with the number of reported partner-inflicted BIs ranging from none to 1,095. Each provided detailed accounts of their IPV exposure and completed ten cognitive tests, scored across sixteen cognitive domains. As cognition is shaped by many factors beyond BI, from years of education to substance use, we measured and accounted for these simultaneously [18,34,35]. This unusually deep characterization allowed us to test how cognition may be affected as partner-inflicted BIs accumulate. We emphasize that what follows concerns partner-inflicted BI and its measurable associations with cognition, not the intelligence or capability of the women who sustained it.

We expressed performance as cognitive age, defined here as the age at which the typical person in the general population performs equivalently. A 30-year-old performing like a typical 48-year-old therefore would have a cognitive age gap of 18 years. Intuitively, some women with severe BI histories were unable to complete particular cognitive tasks, produced scores deemed invalid by the assessment, or both. We therefore excluded their performance from relevant analyses per convention. As a result, the findings we report only capture the performance of survivors whose performance could be validly measured, and we bear in mind that the cognitive losses reported here are likely worse than what current cognitive methods can capture.

## Results

### Two-thirds of women reported at least one partner-inflicted brain injury

We began by quantifying partner-inflicted BI among the women. Two-thirds reported at least one injury (67%; *n* = 139) inflicted by their intimate partner. Over two-fifths (42%; *n* = 86) had multiple partner-inflicted BIs and 27% (*n* = 56) reported at least one BI from strangulation-related alterations in consciousness (Fig. 1 and Table S1). Among injured women, the median was two partner-inflicted BIs, one-quarter reported nine or more, and 15 reported more than 25, with counts reaching as high as 1,095. To limit the influence of extreme counts, we capped injury counts at 25 in our main analysis. This conservative choice did not drive the findings, which remained unchanged when women exceeding the cap were excluded (Table S5).

**Fig. 1.**
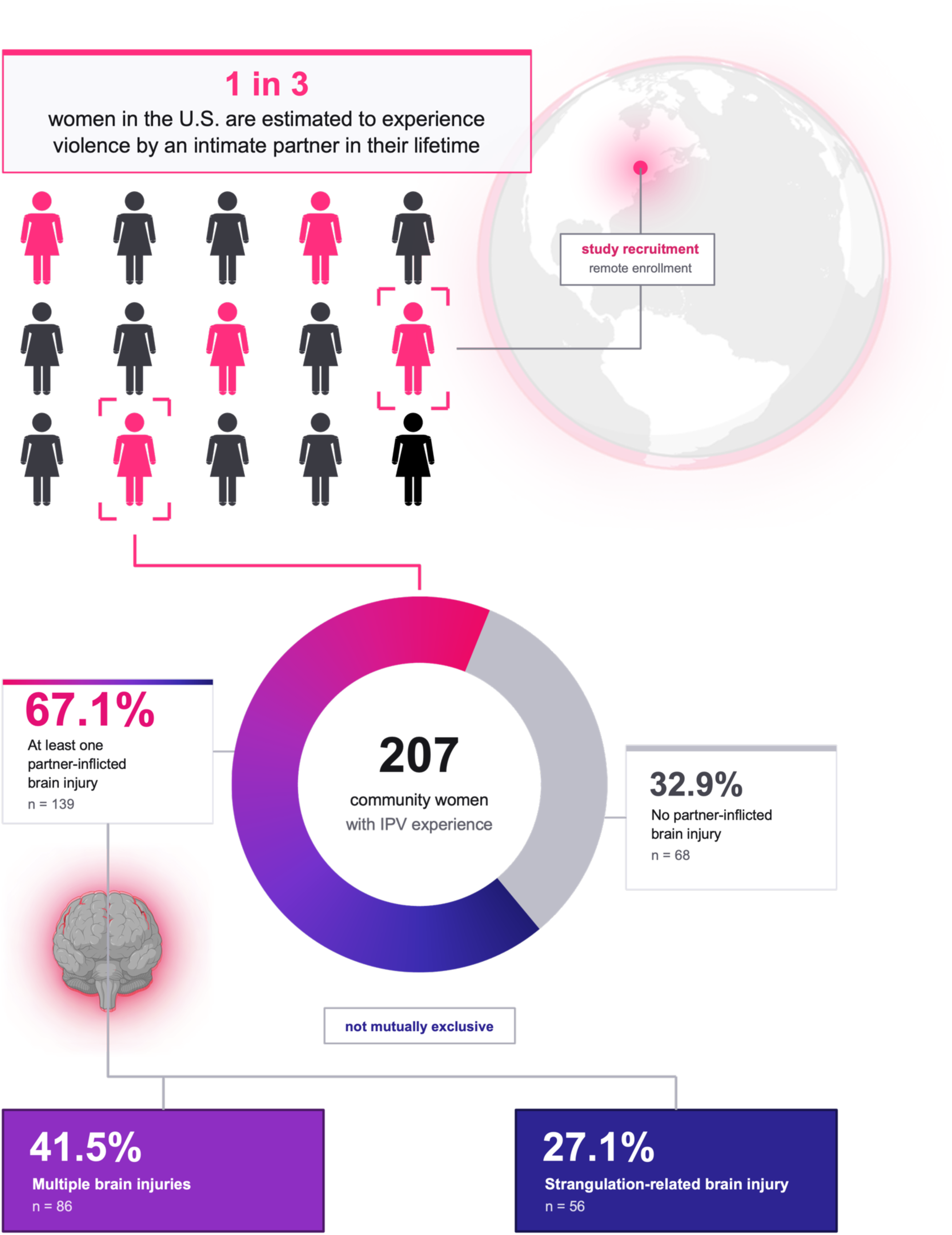
Partner-inflicted brain injury in a community sample of women exposed to intimate partner violence. Over one in three women in the United States are estimated to have experience abuse from an intimate partner in their lifetime. Among the 207 women studied here, all reporting lifetime exposure to partner violence, 139 (67.1%) reported at least one partner-inflicted BI and 68 (32.9%) reported none. Among those who had sustained BIs, 86 women (41.5% of the sample) sustained more than one and 56 (27.1%) sustained a strangulation-related injury; the two categories overlap.

### Cognitive performance fell below population norms across most domains

To establish how these women performed relative to the general population, we next measured cognition across sixteen domains. Scores are age-adjusted, with a population mean of 100 and a standard deviation of 15. Mean scores were below the population mean on 14 of the 16 domains, 11 of which survived Bonferroni correction across sixteen tests (Fig. 2, white hatched bars, and Table S2). Women who had survived any physical violence demonstrated poorer reaction time (mean 88.9; *d* = 0.74; *P_bonf_* < 0.0001), simple attention (90.9; *d* = 0.61; *P_bonf_* < 0.0001), executive function (91.7; *d* = 0.55; *P_bonf_* < 0.0001), cognitive flexibility (92.0; *d* = 0.54; *P_bonf_* < 0.0001), composite memory (92.1; *d* = 0.53; *P_bonf_* = 0.53; *P* < 0.0001), verbal memory (92.6; *d* = 0.49; *P_bonf_* < 0.0001), motor speed (93.2; *d* = 0.45; *P_bonf_* < 0.0001), psychomotor speed (93.5; *d* = 0.44; *P* < 0.0001), global cognition (i.e., the composite measure of overall cognitive function; 93.8; *d* = 0.41; *P_bonf_* < 0.0001), visual memory (94.6; *d* = 0.36; *P_bonf_* < 0.0001) and reasoning (95.1; *d* = 0.32; *P_bonf_* = 0.0003).

**Fig. 2.**
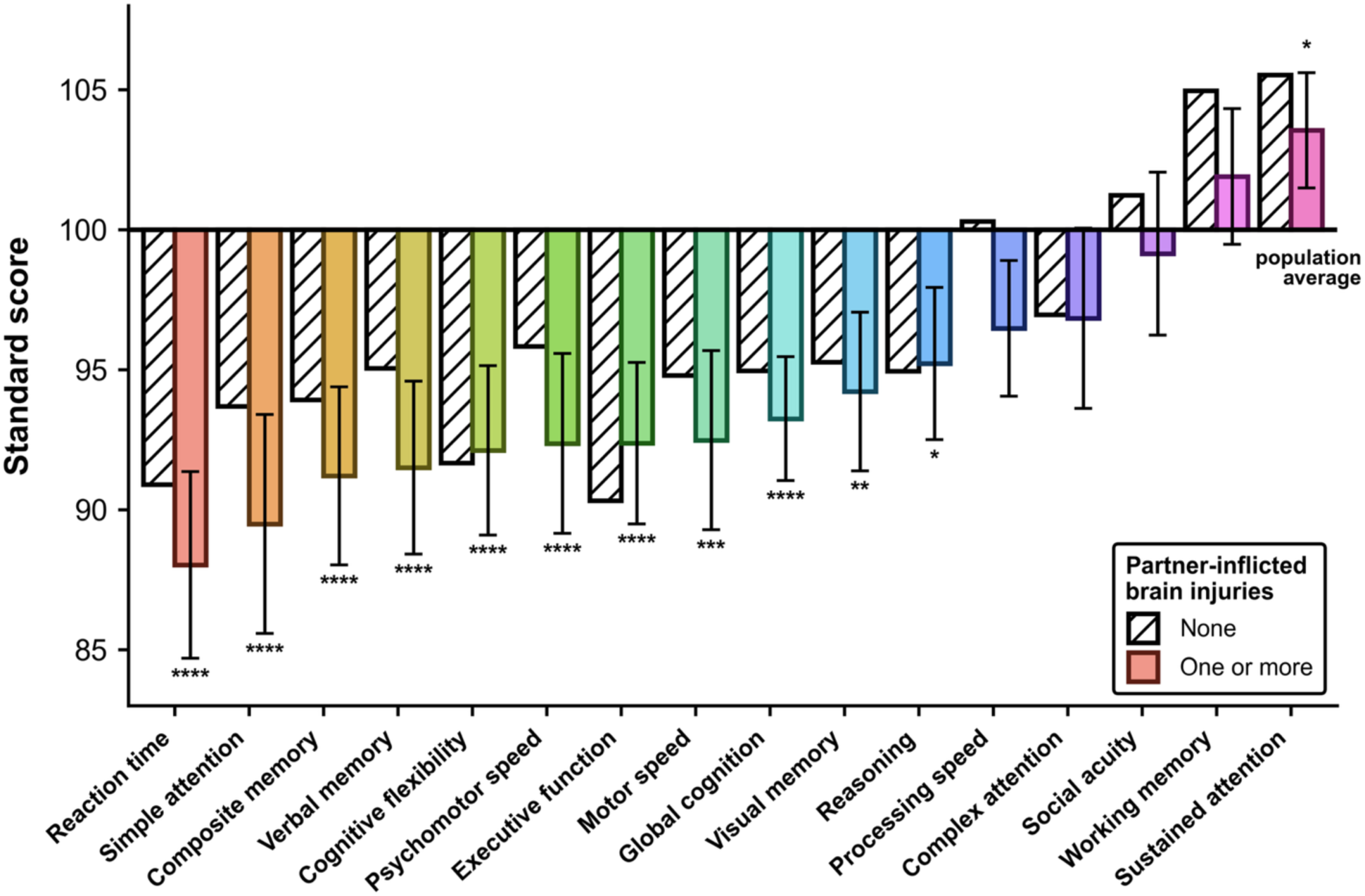
Cognitive performance against population norms in women with and without partner-inflicted brain injury. Bars give the mean standard score for each domain in the 68 women reporting no partner-inflicted brain injury (white, hatched) and the 139 women reporting at least one (coloured). Both groups experienced physical violence from an intimate partner, so the contrast is injury rather than exposure to violence. Domains are ordered by the injured group, lowest to highest, and the horizontal line marks the population average of 100. Error bars are 95% confidence intervals for the injured group. Asterisks denote significance for the injured group against the population average of 100, after Bonferroni correction across sixteen tests: \**P* < 0.05; \*\**P* < 0.01; \*\*\**P* < 0.001; \*\*\*\**P* < 0.0001. The uninjured group is shown for reference only. No significance is marked for it, and no test between the two groups is displayed. However, mean scores fell significantly below the population average on four domains in the uninjured group and significantly below the population average on eleven domains among injured women. Social acuity sits at the population average in both.

We then investigated whether these cognitive deficits were more pronounced in women who reported any history of partner-inflicted BI. The same 14 cognitive domains were below the population mean, with considerably larger deficits among most of them. 11 observed cognitive deficits again remained significant after Bonferroni correction (Fig. 2, coloured bars; Table S2): Women who had sustained at least one partner-inflicted BI demonstrated poorer reaction time (88.0; *d* = 0.80; *P_bonf_* < 0.0001), simple attention (89.5; *d* = 0.70; *P_bonf_* < 0.0001), composite memory (91.2; *d* = 0.59; *P_bonf_* < 0.0001), verbal memory (91.5; *d* = 0.57; *P_bonf_* < 0.0001), cognitive flexibility (92.1; *d* = 0.53; *P_bonf_* < 0.0001), psychomotor speed (92.4; *d* = 0.51; *P_bonf_* = 0.0001), executive function (92.4; *d* = 0.51; *P_bonf_* < 0.0001), motor speed (92.5; *d* = 0.50; *P_bonf_* = 0.0001), global cognition (93.2; *d* = 0.45; *P_bonf_* < 0.0001), visual memory (94.2; *d* = 0.39; *P_bonf_* = 0.001) and reasoning (95.2; *d* = 0.32; *P_bonf_* = 0.011).

Deficits in complex attention and processing speed were smaller and did not survive correction, at 96.9 (*d* = 0.21; *P_bonf_* = 0.269; *P* = 0.017) and 97.7 (*d* = 0.15; *P_bonf_* = 0.620; *P* = 0.039) respectively across the cohort, and at 96.8 (*d* = 0.21; *P_bonf_* = 0.856; *P* = 0.054) and 96.5 (*d* = 0.24; *P_bonf_* = 0.075; *P* = 0.005) among the women reporting at least one partner-inflicted BI. Social acuity, the ability to identify emotional expressions, was indistinguishable from the population mean in both groups, at 99.8 (*d* = 0.01; *P_bonf_* = 1.00) and 99.1 (*d* = 0.06; *P_bonf_* = 1.00).

Mean working memory (102.9; *d* = 0.19; *P_bonf_* = 0.058) and sustained attention (104.2; *d* = 0.28; *P_bonf_* < 0.0001) were above the population mean across the cohort, and remained larger among the women reporting at least one partner-inflicted BI, at 101.9 (*d* = 0.13; *P_bonf_* = 1.00) and 103.5 (*d* = 0.24; *P_bonf_* = 0.015), respectively. Yet, working memory and sustained attention constituted the two domains with the most excluded scores, a point we return to in the discussion.

### Cognitive performance declines as partner-inflicted injuries accumulate

We next examined whether the number of partner-inflicted BIs a woman had sustained predicted her cognitive performance. To express the size of any gradient in interpretable terms, we grouped the women into four bands of injury burden (Methods; Fig. S1). Cognitive scores declined across all sixteen domains as partner-inflicted BI count increased (d = 0.06 to 0.70 from the lowest to the highest band; *P for trend* = 0.0033 to 0.65). The decline remained significant on overall cognitive function, reaction time, psychomotor speed and processing speed following false discovery rate correction (*d* = 0.55 to 0.70; all *q* = 0.032). Overall cognitive function, for instance, fell below the normative mean in each injury burden band, from 95.0 in women reporting none to 87.6 in those reporting ten or more (*d* = 0.30 to 0.82 against the normative mean, all *P* ≤ 0.035; *d* = 0.70 from lowest to highest band, *P* for trend = 0.0065; Fig. 3A, Fig. S2 and Table S3).

**Fig. 3.**
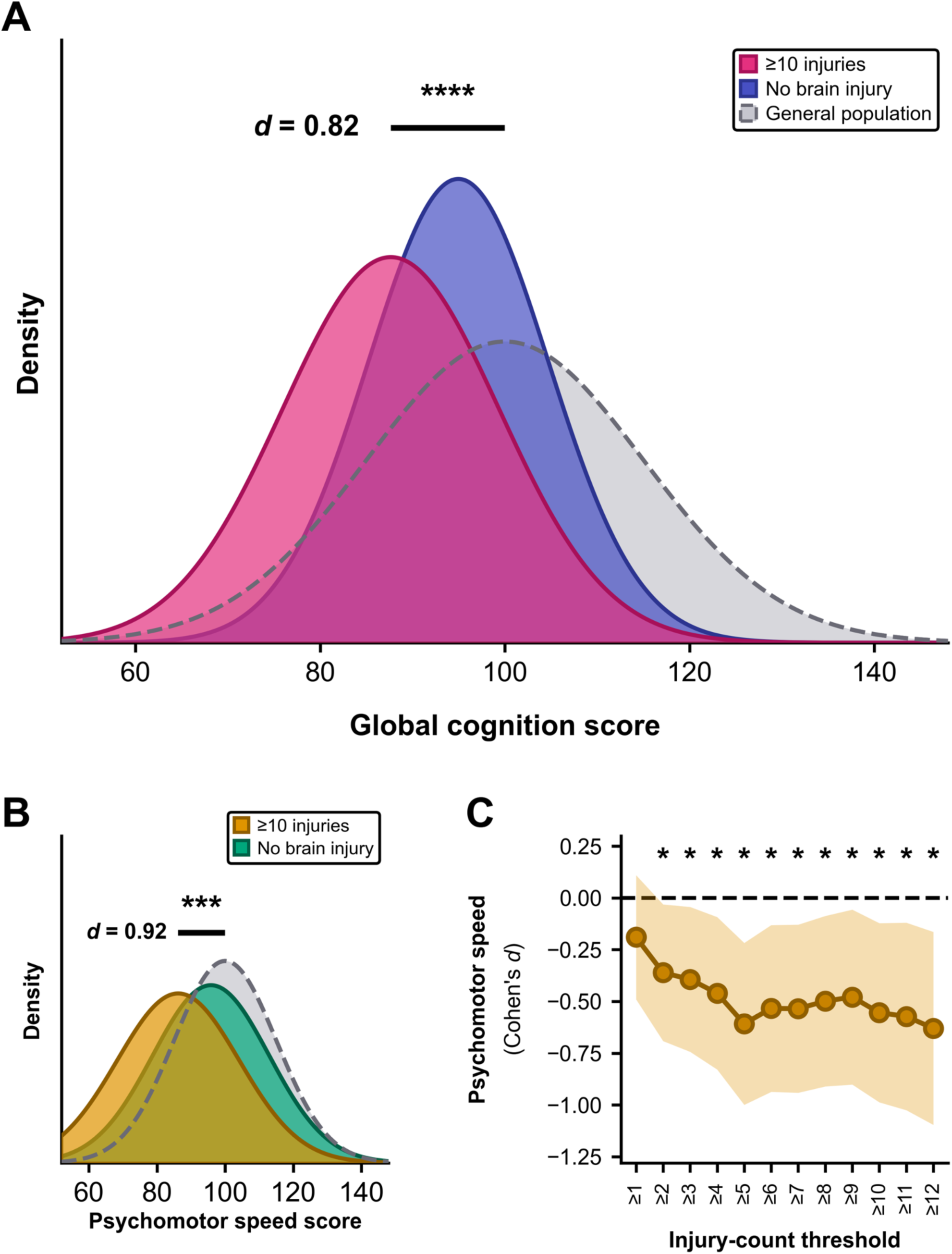
The deficit scales with injury count and is independent of the threshold chosen. **(A)** Global cognition drawn as normal distributions with a common standard deviation of 15, for the general population (grey, dashed, mean 100), women reporting no partner-inflicted BI (indigo, 95.0) and women reporting ten or more (pink, 87.6). The bar marks the difference between the heavily injured group and the population mean, 0.82 SD, *P_bonf_* = 0.0001); the same group sits 0.70 SD below uninjured women in this cohort (d = 0.70, P = 0.007). **(B)** The same construction for psychomotor speed, with women reporting no partner-inflicted BI (green, 95.8) and ten or more (orange, 86.2) against the general population (grey, dashed). The bar marks 0.92 SD below the population mean, *P_bonf_* = 0.002); the within-cohort difference is 0.55 SD (P = 0.015). Asterisks in (A) and (B) denote unadjusted one-sample tests against the population mean (*P < 0.05; **P < 0.01; ***P < 0.001; ****P < 0.0001); the Bonferroni-corrected values are those quoted above and in Table S2. **(C)** The within-cohort contrast on psychomotor speed re-estimated at twelve injury thresholds, each comparing women reporting at least that many injuries against women reporting none. Negative values indicate poorer performance in the exposed group, shading gives the 95% confidence interval, and asterisks mark P < 0.05. Every threshold from two injuries upward yields a significant difference (*d* = 0.36 to 0.63; Table S4); two is the median number of injuries reported by injured women. All sixteen domains are ranked by effect size in Fig. S2.

We then investigated whether these cognitive deficits were more pronounced still in women who reported ten or more partner-inflicted BIs. Deficits were largest in this group, extending to fourteen of the sixteen cognitive domains. Ten observed cognitive deficits remained significant after Bonferroni correction (Fig. 3A; Table S2), namely that women who had sustained ten or more partner-inflicted BIs demonstrated poorer reaction time (79.6; *d* = 1.36; *P_bonf_* = 0.0003), simple attention (85.7; *d* = 0.95; *P_bonf_* = 0.023), psychomotor speed (86.2; *d* = 0.92; *P_bonf_* = 0.002), composite memory (86.9; d = 0.88; *P_bonf_* = 0.018), executive function (87.2; *d* = 0.86; *P_bonf_* = 0.001), cognitive flexibility (87.3; *d* = 0.85; *P_bonf_* = 0.003), global cognition (87.6; *d* = 0.82; *P_bonf_* = 0.0001), verbal memory (87.8; *d* = 0.81; *P_bonf_* = 0.007), motor speed (88.0; *d* = 0.80; *P_bonf_* = 0.010) and processing speed (91.2; *d* = 0.59; *P_bonf_* = 0.016). Women in this greatest injury burden group scored on average 87.6 on overall cognitive function, therefore performing worse than nearly 80% of the general population (*d* = 0.82, P < 0.0001).

A deficit relative to population norms could reflect the conditions of living with a physically abusive partner, such as chronic stress or disrupted sleep, rather than brain injury. To mitigate this possibility, we compared women who had sustained a partner-inflicted BI against women in the same cohort who had endured physical partner violence with no history of partner-inflicted BI. Women reporting ten or more injuries scored below their uninjured counterparts on all sixteen domains (*d* = 0.06 to 0.70; *P* = 0.006 to 0.76; Table S3). The largest differences were on global cognition (*d* = 0.70; *P* = 0.007), reaction time (*d* = 0.64; *P* = 0.014), processing speed (*d* = 0.55; *P* = 0.006) and psychomotor speed (*d* = 0.55; *P* = 0.015), the same four domains the full-sample analysis above identified (Fig. 3A, Fig. S2 and Table S3).

Although we defined the most severe exposure group as ten or more BIs, this cut-off is one of many possible. To test whether our finding depended on our selected cut-off, we repeated the comparison at every threshold from one to twelve injuries (see Methods). A threshold of one injury separates the presence of injury from its absence rather than grading severity, and is treated separately below. From two injuries upward, which was the median number reported by injured women, cognitive scores were lower in the exposed group at every threshold on 13 of the sixteen cognitive domains (*d* = 0.01 to 0.73). For instance, psychomotor speed and processing speed were significantly lower at every one of those thresholds (*d* = 0.36 to 0.63; all *P* ≤ 0.032; Fig. 3C and Table S4). On global cognition, the deficit also worsened as the threshold rose, reaching significance from four upward and becoming nearly five times worse between one or more BIs (*d* = 0.15; *P* = 0.330) and twelve or more (*d* = 0.73; *P* = 0.011).

Finally, women who experience IPV often are assessed by whether they have a history of brain injury rather than by how many they have sustained [36,37]. To test whether injury presence alone was informative, we compared survivors reporting any partner-inflicted BI with those reporting none. The cognitive differences were small and imprecise (d = 0.01 to 0.26, all P ≥ 0.13; the largest, on working memory, 95% CI [−0.09, 0.61]), and therefore too uncertain to statistically separate a woman who has sustained a BI from one who has not.

Many factors beyond partner-inflicted BI shape cognition, including schooling, psychiatric distress, and injuries sustained before the relationship. We therefore adjusted every domain simultaneously for these potential confounders among eight others. After adjustment, injury count was associated with poorer performance on fourteen of the sixteen domains, including six meeting conventional levels of statistical significance at *P* < 0.05 versus 0.8 expected by chance (Table 1). The strongest was psychomotor speed (partial *r* = −0.22, *P* = 0.003), followed by reaction time, processing speed, motor speed, global cognition and verbal memory (partial *r* = −0.14 to −0.20, all *P* < 0.05). The consistency of direction across domains was itself unlikely under the null (two-sided sign test *P* = 0.004), though the domains share participants and a common exposure and so are not independent tests.

**Table 1.**
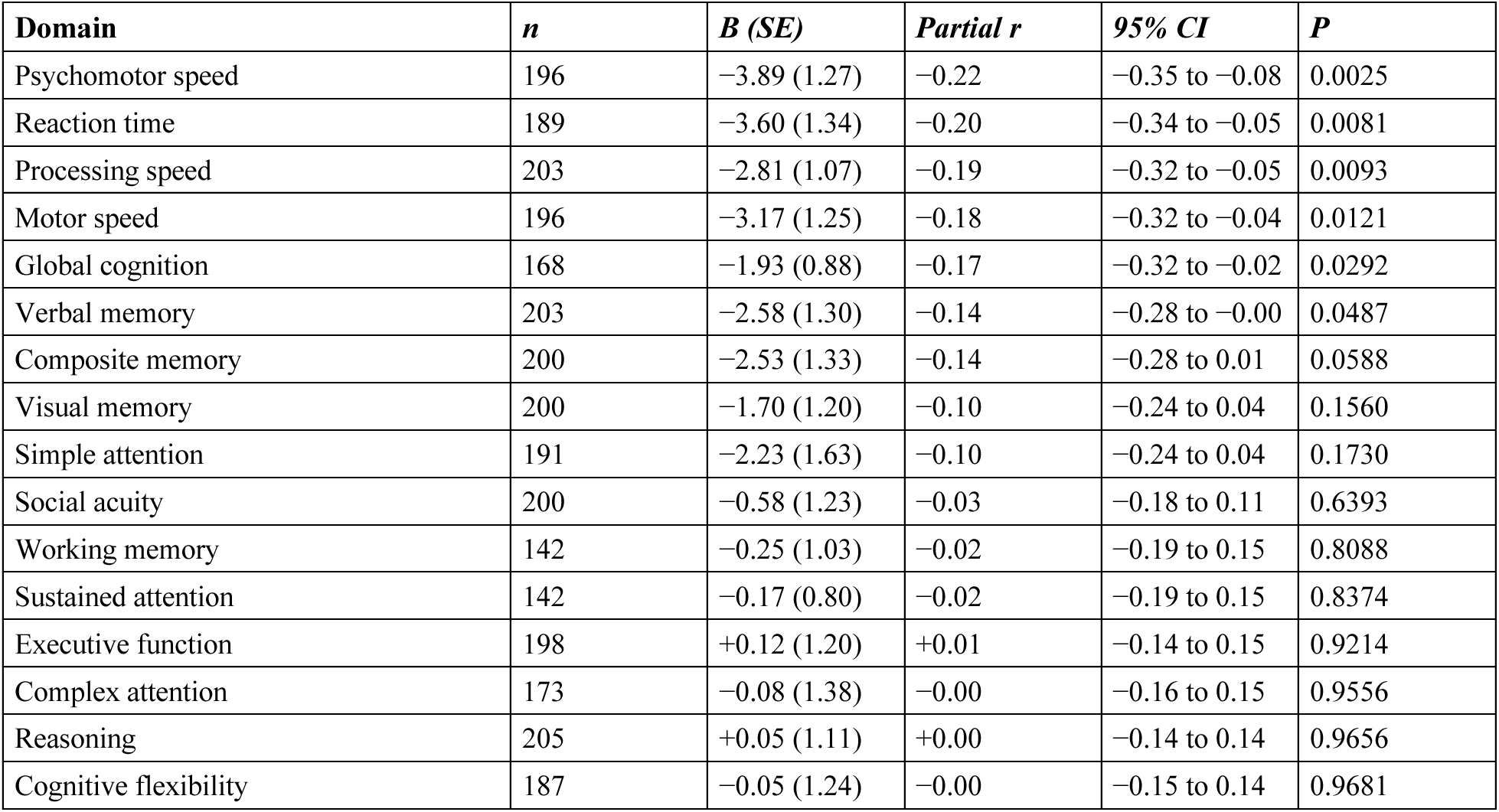
Adjusted associations between cumulative partner-inflicted brain injury and cognitive performance. Multivariable linear regression of standard score on log injury count, adjusted for age, education, race, non-partner adult and childhood brain injury, post-traumatic stress, depression, anxiety, alcohol use, childhood maltreatment, and abuse severity. *B* is the unstandardized coefficient with its standard error, giving the change in standard score per unit increase in log injury count, and *P* the two-sided probability. All sixteen domains are reported, from a single specification applied to each in turn. Six associations reach *P* < 0.05, against 0.8 expected were no association present, and fourteen of the sixteen coefficients are negative (two-sided sign test *P* = 0.004). Under Benjamini-Hochberg control across the sixteen domains, four remain significant at *q* < 0.05 (psychomotor speed, reaction time, processing speed and motor speed).

We recognized that asking women whose memory may have been impaired to recall how many injuries they sustained creates an inherent tension, since those with the most injuries might remember the fewest. We therefore tested whether memory predicted reported count. Women with poorer memory reported slightly more injuries, not fewer (composite memory *r* = −0.13, *P* = 0.07; verbal memory −0.14, *P* = 0.05), the opposite of what one might expect with forgetting or poor memory. Speed domains were associated with injury count more strongly than memory domains, the reverse of the pattern memory-driven under-reporting would produce (Table S10).

### Cumulative brain injury corresponds to more than a decade of additional cognitive aging

To convey the magnitude of these deficits in more intuitive terms, we translated the already observed standardized deficits into cognitive age, defined as the age at which the typical person performs at the same level. We derived cognitive age separately for each domain with a raw score that changes with age. Given that global cognition is an average of age-standardized scores across multiple cognitive domains, and therefore has no raw performance measure that changes with age, a cognitive age for global cognition cannot be calculated.

Women reporting ten or more partner-inflicted BIs performed over two decades older than the general population across eight cognitive domains (20.8 to 28.1 years; Table S12): simple attention (28.1 years; *P* = 0.0002), reaction time (27.8; *P* < 0.0001), verbal memory (26.0; *P* < 0.0001), motor speed (24.6; *P* < 0.0001), composite memory (22.7; *P* < 0.0001), cognitive flexibility (22.6; *P* < 0.0001), executive function (21.9; *P* < 0.0001) and psychomotor speed (20.8; *P* < 0.0001); see Fig. 4A for an illustrative example. Across the eleven domains significantly older than the general population, gaps ranged from 11.4 to 28.1 years.

**Fig. 4.**
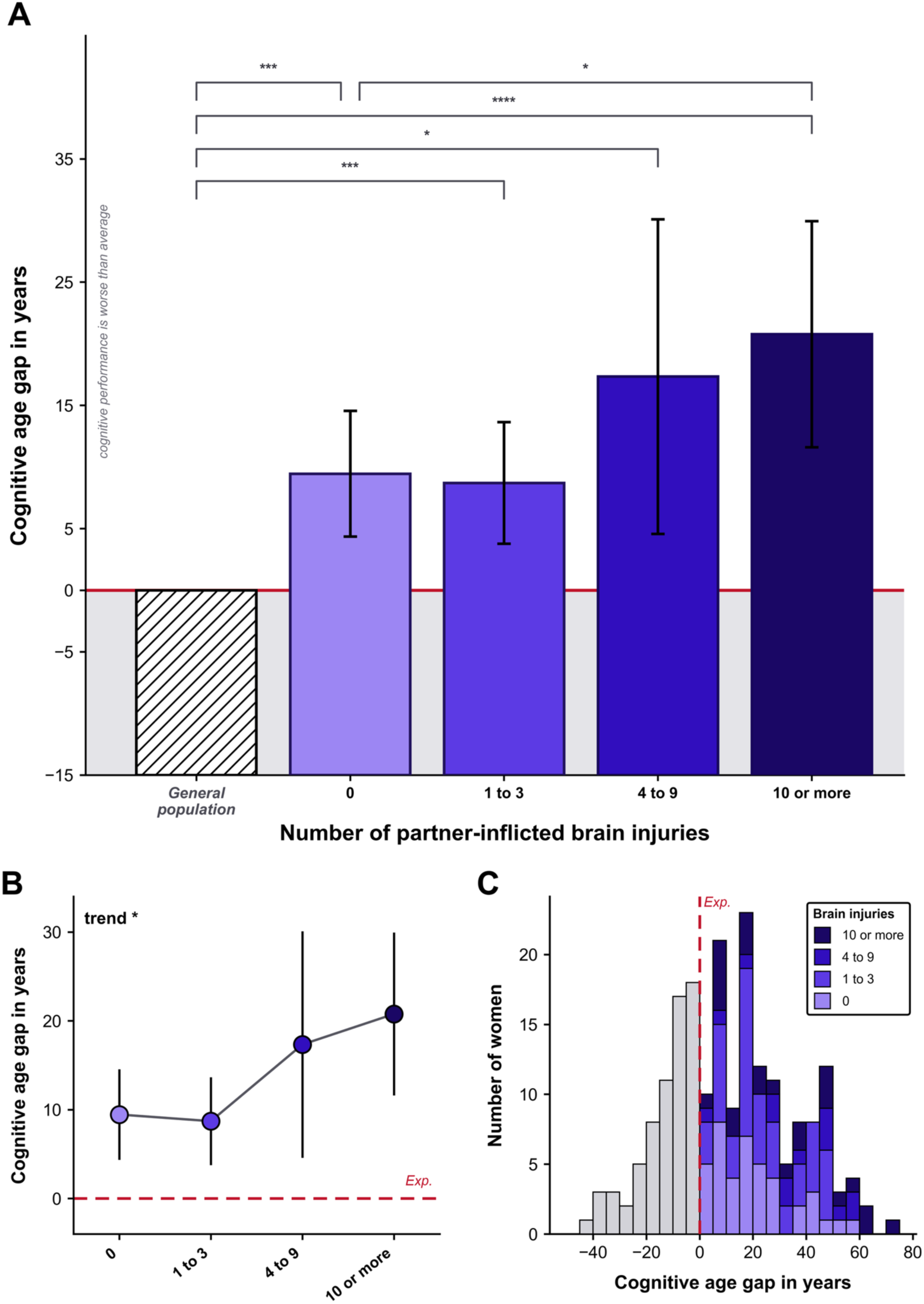
Cognitive age gap by cumulative partner-inflicted brain injury. **(A)** Mean cognitive age gap in years, derived from psychomotor speed, for each exposure band, against a general-population reference of zero (hatched bar). Women reporting no partner-inflicted brain injury sat 9.5 years above the reference, against 8.7 years for one to three injuries, 17.3 for four to nine, and 20.8 for ten or more (n = 63, 80, 22 and 32 with a computable gap). Error bars are 95% confidence intervals. Brackets give tests against the population reference, and the lowest pair the within-cohort comparison between women reporting none and women reporting ten or more, 11.3 years (P = 0.033). Shading below zero denotes performance better than the population average for age. **(B)** Band means with 95% confidence intervals across the four ordered bands (P for trend = 0.0122). **(C)** Distribution of individual cognitive age gaps, stacked by exposure band. Bars left of the dashed line denote women performing at a cognitive age younger than their own. The proportion performing better than expected for their age fell from 35% among women reporting no injury to 22% among those reporting ten or more, against 50% expected in the normative sample. Asterisks: *P < 0.05; **P < 0.01; ***P < 0.001; ****P < 0.0001.

Comparing women who had sustained ten or more partner-inflicted BIs against survivors who had sustained no BI, they performed over a decade older on four cognitive domains: simple attention (14.2 years; *d* = 0.40, *P* = 0.074), verbal memory (13.7; *d* = 0.44, *P* = 0.036), reaction time (11.9; *d* = 0.51, *P* = 0.036) and psychomotor speed (11.3; *d* = 0.51, *P* = 0.033). The difference reached significance on the last three of these and on processing speed (8.6 years; *d* = 0.52, *P* = 0.024; Table S12).

The gap widened as a woman sustained more partner-inflicted BIs (Fig. 4A and 4B). On psychomotor speed, for instance, even sustaining two or more partner-inflicted BIs equated to at least one decade, and potentially over two, of additional cognitive age relative to the general population (16.4 years; 95% CI [11.1, 21.8], *P* < 0.0001). The gap rose to 20.8 years in those reporting ten or more, across a gradient spanning the four exposure bands (*P* for trend = 0.0122). Treated as a continuous exposure rather than in bands, each log-unit increase in BI count corresponded to 4.58 additional years of cognitive age, adjusted for the same eleven covariates (SE 1.60, *P* = 0.005).

Calculating cognitive age can also produce negative values, meaning that some women performed better than expected for their chronological age (Fig. 4C). This is to be expected as roughly 50% of individuals should perform better than their predicted cognitive age within a normal population. Yet only about a third of survivors of physical IPV (35%, *n* = 68 of 197) performed better, whether or not they had sustained a partner-inflicted BI, falling to about one in five (22%, *n* = 7 of 32) among those who had sustained at least ten BIs from an intimate partner.

To determine whether the association depended on the specific analytic decisions we had made, we re-estimated it using alternative injury definitions, band schemes, outcome scaling, and covariate sets. The association held throughout (Tables S4 to S7). Finally, we asked whether the association reflected one mechanism of injury rather than cumulative injury itself, since blows to the head and strangulation injure the brain by different routes, the first by force and the second by depriving it of oxygen. The association was carried by blows to the head, although far fewer strangulation injuries were reported and the comparison is correspondingly less well powered to detect an effect of that mechanism (Table S11).

### Validity criteria exclude the most impaired women from the analysis

Every result reported above follows the standard practice of discarding cognitive scores that the assessment platform flags as invalid. To determine whether we were disproportionately excluding data from women with poorer cognitive performance, we compared the cognitive performance of women with at least one invalid score with those whose scores were never flagged.

The cognitive assessment platform flags a score as invalid when a response of a participant appears incompatible with genuine effort, and standard practice is to discard it. Forty percent of the women (40%; *n* = 82) in our study had at least one score flagged and accordingly excluded for the respective cognitive measures. In particular, women were consistently getting their scores flagged and excluded on two cognitive domains: their ability to stay focused on a task for an extended period of time (31%; *n* = 64) and their ability to hold information in their mind while performing a task, known as working memory (31%; *n* = 64; Fig. 5A).

**Fig. 5.**
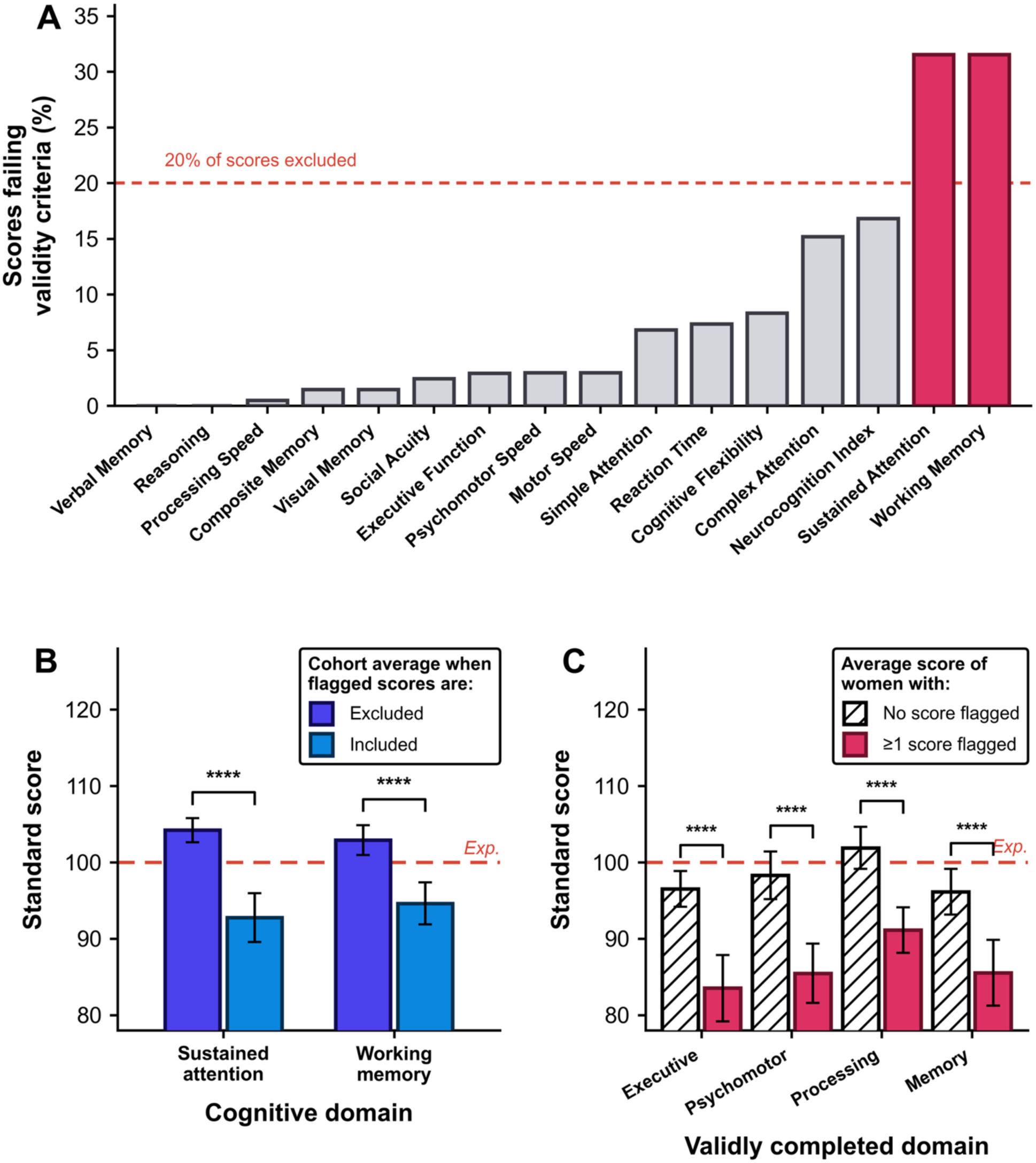
Excluding test scores that fail validity criteria removes the most impaired women. Scores flagged by the cognitive battery’s embedded validity indicator are excluded from all analyses reported here. Scores are flagged as potentially unreliable based on response speed, error patterns, or other indicators of lack of genuine effort. **(A)** Percentage of scores flagged, by domain. Sustained attention and working memory, both scored from the four-part continuous performance test, are flagged in 31% of scores each (pink); the dashed line marks 20%. **(B)** Cohort average on those two domains under two conventions, with flagged scores excluded (indigo) or included 17 (blue). Excluding them places both domains above the population average of 100; including them places both below. **(C)** Average score of women who had at least one score flagged elsewhere in the battery (pink) against women who had none (hatched), shown only for domains the battery accepted from both groups. Women with a flagged score scored 11 to 13 points lower on the domains they did complete validly. Bars give 95% confidence intervals. Asterisks: *P < 0.05; **P < 0.01; ***P < 0.001; ****P < 0.0001.

When their flagged scores are excluded, survivors of physical partner violence appeared to have better sustained attention and working memory than the general population, scoring three to four points above the normative mean of 100 (*d* = 0.28 and 0.19; *P* < 0.0001 and *P* = 0.004). Yet, had we included the flagged scores in our original analysis, the survivors would have appeared to perform measurably worse than the general population (*d* = 0.48 and 0.36; *P* < 0.0001 and *P* = 0.0001) (Fig. 5B). When comparing women who had at least one flagged score with those who had none, on the domains the test did accept from them, such as their speed of thinking and their memory, those with a flagged score performed 11 to 13 points lower (Fig. 5C). In other words, the scores being excluded belong to the women with the most severe cognitive deficits in other domains.

## Discussion

Partner-inflicted BI is among the most common form of head trauma and among the least characterized in its impacts [18–20,38]. In the largest cognitively assessed cohort of survivors of physical IPV, women reporting ten or more injuries performed at a cognitive age more than two decades above the general population and over a decade above BI-free women also exposed to partner violence. Women with any BI performed significantly lower than the general population on a variety of cognitive measures. We consider what a deficit of this size means for survivors, why it may exceed established estimates, what our measurements miss, and what the findings imply for clinics and courts.

### A cognitive deficit of this size is consequential in ordinary life

We found that women who had suffered ten or more partner-inflicted BIs performed at a cognitive age over two decades older than the general population, and over a decade older than women who had endured physical IPV without sustaining a BI. In practical terms: the average 30-year-old woman who has sustained ten or more BIs from her partner performs cognitively on psychomotor speed as a 51-year-old woman in the general population, likely while having to navigate employment and childcare as well as possible legal demands, and continuing efforts to remain safe.

Performing cognitively at least a decade older than expected is demonstrably concerning. Notwithstanding how poorer cognitive ability is associated with earlier loss of independence [25], more motor vehicle crashes [39], and the ability to be employed [40], this result is particularly grave because partner-inflicted injuries may compromise the very cognitive capacities women must rely on to rebuild an independent life. These cognitive abilities, especially when significantly impaired, may compromise a survivor’s ability to leave abusive relationships, attain medication, care for their children, sustain employment, navigate legal proceedings, and remain safe post-violence.

Social acuity, the ability to distinguish emotional expressions, was the only cognitive domain in which performance was indistinguishable from the general population (*d* = 0.01, *P* = 1.00), and it did not decline detectably as injury burden rose (*r* = −0.05, *P* = 0.52). This relative preservation may be particularly meaningful in the context of an abusive relationship. Detecting subtle changes in a partner’s facial expression may allow a woman to recognize anger or agitation, anticipate escalating violence, and take steps to protect herself. Repeated exposure to interpersonal threat could therefore preserve, or potentially sharpen, attention to socially relevant emotional cues even as other cognitive abilities deteriorate as partner-inflicted BIs accumulate.

We cannot determine whether this finding reflects adaptation to chronic threat or coincidence, but the cognitive capacity most relevant to detecting interpersonal threat was the cognitive that appeared to remain intact.

### Counting brain injuries helps detect cognitive harm

The present study demonstrates that cognitive deficit becomes more pronounced as partner-inflicted BIs accumulate. Women with any partner-inflicted BI did not differ significantly from women with none, while cognitive performance worsened with increasing injury burden. Prior studies point in a similar direction. Greater BI severity has been associated with poorer memory, learning, and cognitive flexibility [41], injury count with poorer visual memory [42], and repetitive head injury with greater cognitive symptom burden [43]. A recent study also found that learning and delayed-recall deficits were most apparent among women reporting more than six IPV-BIs [44]. Together, these findings suggest that cumulative exposure is important for understanding cognitive outcomes after partner-inflicted BI.

Our study extends this literature by showing that injury count is informative across a broad cognitive profile and remains so across multiple ways of defining exposure. This has a methodological implication for future studies. Classifying survivors only by the presence of BI may therefore obscure substantial variation in cumulative exposure and weaken associations with cognition. Alongside evidence that brain injury is often repetitive in IPV [45], our findings suggest that injury count should be treated as a core dimension of exposure to better capture variation in cognitive outcomes.

### A well-studied deficit in an understudied population

BI in sports and the military provides the firmest available benchmark against which to place the present findings. Repeated head injury in athletes and veterans has been studied extensively, attracting billions of dollars in funding and dedicated federal programs [32], and more than four hundred articles on football concussion in the New York Times between 2007 and 2013 alone [38].

Placing these deficits in context is difficult, because effect estimates drawn from different batteries, reference groups, exposure definitions and intervals since injury are not strictly comparable. Provided this caveat, meta-analyses report cognitive deficits of 0.29–0.49 standard deviations after sports concussion and 0.12–0.33 after blast-related mild traumatic BI [46–48], with poorer performance also observed after repetitive subconcussive exposure and recurrent concussion [49,50]. Here, women reporting partner-inflicted BI showed deficits of 0.45 standard deviations in global cognition, 0.51 in psychomotor speed, 0.59 in memory and 0.80 in reaction time. These estimates sit at or above those reported in sports and military populations. Similarly, a 30-year study associated two or more head injuries with cognitive decline equivalent to 9.7 additional years of age, compared with the 16.4-year cross-sectional cognitive-age gap observed here [51].

Importantly, prior sports literature has characterized athletes with three or more prior concussions as a highly exposed group [52], whereas here, the median injured woman reported two BIs, a quarter reported nine or more, and counts reached 1,095. In our highest band of ten or more BIs, the deficit reaches 0.82 standard deviations below the population and 20.8 years of cognitive aging. No published benchmark exists at that level of exposure, and we report these estimates to describe a range that has not previously been documented.

A variety of factors may explain the comparatively large cognitive deficits observed here, including the circumstances surrounding each BI. Athletes, for instance, wear protective equipment manufactured to standardised impact criteria [53]. When an injury is suspected, they are removed from play, examined by trained staff, and given a defined interval before returning [54]. Even there, quantifying cumulative head-impact exposure remains methodologically unresolved [55]. These protections did not extend to the present cohort, as seldom do IPV survivors receive immediate assessment [41], have their injuries monitored [20,56], have any opportunity to remove themselves from the setting in which the injury occurred, likely recover before another injury is sustained [18,19], or wear protective headgear at the time of partner abuse. Help-seeking following BI is itself additionally constrained by stigma, unmet service needs, and mandatory reporting requirements [57–59].

### The true cognitive harm is likely larger

We anticipate that the real cognitive harms of partner-inflicted BI are worse than reported here. The women who entered were safe enough to be contacted, had time and privacy to participate, and retained sufficient cognitive capacity to complete detailed interviews, cognitive testing, and be coherent enough to communicate. The cohort was also unusually educated with 62% holding a college degree against 40.1% of women aged 25 and over nationally [60]. Because higher education predicts stronger cognitive performance and may itself protect against partner violence [61,62], this sample was both less representative of women at greatest risk and more likely to score above population norms.

In addition, among the women for whom the timing of injury was recorded, the most recent partner-inflicted BI preceded testing by a median of 7.5 years, and 94% had been injured more than a year earlier. Cognitive performance can improve after BI, particularly in the first year following severe injury [63], so testing years later may capture the impairment remaining after recovery rather than its full initial severity. Women still living under immediate threat, or whose injuries left them unable to complete a research visit, are less likely to participate in our studies, as well as women residing in shelters.

The most severe form of bias here is survival itself. By definition, this study could include only women who lived following their BI. Put another way, those most likely to have been cognitively affected by partner-inflicted BIs were also the most likely to have been unable to participate in our study or to not have survived.

Our analysis choices were also conservative in ways that likely understate the harm. We capped injury counts at 25 even though the highest reported count was 1,095, excluded scores flagged by the cognitive battery [64–69] even though they belonged to women who performed worse on accepted domains, and restricted cognitive ages to 18 to 90 years, meaning that performance below the oldest reference group could not make the estimated gap any larger. Each decision compressed the most severe exposure or impairment, making the cognitive harms observed here more likely to be underestimated than overstated.

### Interpretation and limits

As a cross-sectional study, we have no pre-injury cognitive baseline. We therefore cannot establish that partner-inflicted brain injuries caused the observed deficits, nor rule out that the women who went on to sustain multiple injuries were already performing poorly on cognitive testing. Yet, we have strong reason to assume causality. In other populations, head injury has been shown to precede cognitive decline rather than follow it in a community cohort followed for thirty years [51]. Moreover, cumulative head-impact exposure across a career in contact sport is associated with later cognitive impairments [70], and with chronic traumatic encephalopathy at autopsy, whose severity tracks dementia diagnosed before death [71,72]. These designs, however, cannot establish temporal order. Encouragingly, efforts to quantify the long-term cognitive risk carried by IPV are beginning [73].

In addition, cognition was measured once, with a single battery, and the highest-exposure bands contain relatively few women, which is reflected in the width of their confidence intervals. It also remains an open question how to quantify a survivor’s cognitive impairment when the deficit is so severe that the survivor cannot complete the cognitive assessment task itself [74].

Finally, we translated cognitive performance to cognitive age so that the finding could be interpreted by a clinician, a court, or a legislator. However, we acknowledge that a difference in years describes how a younger woman resembles an older one and does not establish that her brain itself has aged.

### Making an invisible trauma visible in clinics and courts

Screening for partner violence typically assesses physical injury and psychological distress, but rarely cognition [20]. In the context of deficits equivalent to more than a decade of cognitive aging, omitting cognition risks missing a potentially major consequence of partner-inflicted BI. Screening for BI count is therefore vital for better capturing the cognitive harms of IPV.

Detection matters because effective cognitive treatment exists. Cognitive rehabilitation exists that targets memory, attention, processing speed, and executive function, while occupational and speech-language therapies help women apply compensatory strategies [75]. Cognitive behavioral therapy adapted for BI can also treat co-occurring anxiety and depression when delivered with manuals tailored to BI victims [76]. These treatments should be routinely integrated into care for partner violence. Likewise, in sports and military medicine, an identified BI initiates established pathways for monitoring, rehabilitation, and graded return to activity [54,77]. Women injured by intimate partners would benefit from the same continuity of care, particularly as BI does not invariably heal steadily, and some survivors deteriorate years later [78]. A single cognitive assessment or treatment should therefore not be followed by an assumption of recovery.

Of serious importance are the legal implications of severe cognitive deficits. Our findings suggest that BI, especially when repeated, can severely affect the very capacities on which legal judgments depend. The impaired memory observed here may make the chronology of repeated violence less precise, while slowed processing and executive dysfunction commonplace in our cohort may impede testimony, paperwork, and compliance with legal procedure. If these consequences are interpreted as dishonesty, instability, or noncompliance, a woman can be punished for the injury her partner caused. Courts and disability evaluators should not automatically discard cognitive scores flagged as unreliable. Here, women with flagged scores performed worse on measures the same battery accepted, indicating a need for fuller evaluation rather than erasure from the record. BI-informed assessments and procedural accommodations are therefore necessary in disability, criminal, and custody proceedings, particularly where cognitive impairment might otherwise be used to question a woman’s credibility or fitness to be a custodial parent [56].

We note that translating newly established harms of partner violence into legal recognition has clear precedent. After non-fatal strangulation was linked to roughly six- to sevenfold higher odds of attempted and completed intimate partner homicide [79], federal law made strangulation of an intimate partner a felony within federal jurisdiction in 2013 [80], and by 2023 all 50 states had criminalised non-fatal strangulation as a felony, Ohio being the last to do so [81]. The substantial cognitive harms documented here strongly suggest a comparable legal response, with the law recognizing repeated partner-inflicted BI as a grave and cumulative consequence of IPV.

The brain injuries endured by survivors, and their cognitive harms, often remain invisible because clinics and courts rarely look for them. Here, we offer a metric of cognitive age that quantifies one of the many serious harms of partner-inflicted violence. In doing so, we hope to advance meaningful efforts to measure the harms survivors have immeasurably endured.

## Data Availability

The de-identified data supporting this study are available from the corresponding author upon reasonable request. All data will be uploaded to the Federal Interagency Traumatic Brain Injury Research (FITBIR) data repository after completion of the study, at https://fitbir.nih.gov/.

## Acknowledgments

We sincerely thank the women who took part in this study, and the study staff who contributed to data collection and curation. We also thank Harvard Catalyst for biostatistics consultation on the statistical analyses conducted in this study. CWM was supported by the Harvard-UK Fellowship and Clarendon Fund during research.

## Author contributions

Conceptualization: CWM, EMV. Methodology: CWM. Data analysis: CWM. Data curation: EMV. Visualization: CWM. Funding acquisition: EMV. Supervision: EMV. Writing of original draft: CWM. Reviewing and editing: CWM, EMV.

## Data, code, and materials availability

Direct identifiers have been removed from the analysis dataset. The de-identified data supporting this study are available from the corresponding author upon reasonable request. All data will be uploaded to the Federal Interagency Traumatic Brain Injury Research (FITBIR) data repository after completion of the study, at https://fitbir.nih.gov/.

## Supplementary Materials

### Materials and Methods

#### Experimental design

This was a cross-sectional study of cognitive functioning in community-dwelling women with lifetime exposure to physical intimate partner violence. The study was designed to test whether cognitive performance declines as partner-inflicted injuries accumulate.

#### Participants and recruitment

Data collection took place between 2020 and 2026 as part of a larger ongoing study of partner-violence-related brain injury. Participants were recruited through flyers distributed by community partners, an institutional online research platform, and social media. Community partners included hospital-based programs, police-department-affiliated initiatives, and community organizations focused on violence prevention and survivor support. To be eligible, women had to have experienced at least one instance of physical partner violence and be at least 18 years old. A personal history of brain injury was not required. Of 210 eligible women with both brain injury and cognitive data, three were later excluded: two for pre-existing neurological conditions requiring neurosurgery, and one for inconsistent self-report across instruments, leaving 207. All processes and procedures were approved by our hospital’s Institutional Review Board (IRB), and every participant provided informed consent.

#### Procedure

Sessions were conducted by video conference and completed in a single session. Participants were instructed to find a private, distraction-free location. Those without a personal computer were offered a study device. Following informed consent, demographic information was collected by interview. Participants then completed the computerized cognitive battery independently before the abuse and injury interviews. Study staff offered participants the opportunity to prepare a safety plan in advance, including strategies for discreetly pausing the session. Participants could take breaks or withdraw at any point and received a gift card on completion.

#### Exposure assessment

Brain injury exposure was assessed with two structured interviews: the Brain Injury Severity Assessment (BISA) and the Ohio State University Traumatic Brain Injury Identification Method (OSU TBI-ID). The BISA applies the criteria of the American Congress of Rehabilitation Medicine [82] to anything a partner did to the woman, defining a brain injury as a traumatically induced physiological disruption of brain function manifested by loss of consciousness, loss of memory for events immediately before or after the event, alteration of mental state at the time, or focal neurological deficit. The OSU TBI-ID documents lifetime injuries from any cause and asks about loss of consciousness, being dazed, or a gap in memory. A brain injury identified by either interview was counted. Following this definition and prior work in this cohort [42,83–85], alterations in consciousness resulting from violent shaking and from strangulation-induced anoxia or hypoxia were also classified as brain injuries.

Where a participant reported an alteration in consciousness following an incident with a partner, follow-up questions established the mechanism. Events attributed to strangulation or choking were classified as anoxic; events attributed to traumatic force to the head were classified as blunt-force trauma. Strangulation or shaking events that did not produce an alteration in consciousness were not recorded.

The primary exposure was the total count of partner-inflicted brain injuries: blunt-force, anoxic, the two combined categories, violent shaking, and moderate-to-severe events. Counts were capped at 25, beyond which an accurate estimate is difficult to obtain, following prior work in this cohort [42]. Fifteen women, 11% of those exposed, exceeded the cap, with counts up to 1,095. Capping treats these women identically to a woman reporting 25 events and is therefore conservative. A mild-only definition excluding moderate-to-severe events is reported as a sensitivity analysis (Table S7). Counts were log-transformed as log(1 + count) for continuous analyses. Injuries sustained outside the relationship, in adulthood and in childhood, were documented separately and retained as covariates.

#### Cognitive assessment

Cognition was measured with CNS Vital Signs, a computerized battery yielding age-adjusted standard scores with a population mean of 100 and standard deviation of 15 across the sixteen domains reported by the version of the battery administered here [86]. Domain names follow the battery’s own reporting, with one exception. The domain reported here as global cognition is the battery’s Neurocognition Index, the average of its five core domain scores: composite memory, psychomotor speed, reaction time, complex attention and cognitive flexibility. The battery was chosen because it offers a comprehensive assessment of cognitive performance while remaining accessible to a population that is difficult to reach. Administration and scoring are automated, so performance does not depend on rater judgement, which matters where an examiner aware of a woman’s abuse history might otherwise influence testing. Its subtests are computerized adaptations of conventional neuropsychological tests, including the Stroop, Shifting Attention, Continuous Performance and Symbol Digit Coding tasks (Table S9), and concurrent validity against their standard forms has been established [86]. It can be delivered without a laboratory visit, which allows assessment of women for whom in-person testing is impractical or unsafe. All cognitive assessments were administered remotely, in a private location of the participant’s choosing, with a member of the study team available by video for the duration. The normative sample for this battery was tested under supervised administration. The battery reports raw scores alongside age-adjusted standard scores, so the normative relation between age and raw performance can be estimated directly from the data. The battery applies an embedded validity indicator to each domain and flags scores it judges unreliable [64], and the consequences of deleting those scores are examined in the main text and in supplementary text.

#### Derivation of cognitive age

Cognitive aging was estimated by calculating how many years older the average woman in the general population would need to be to show the same performance on a given measure. Because the battery reports both raw and age-adjusted standard scores, the two coefficients needed for this conversion can be estimated by regressing raw score on age and standard score. The age coefficient gives the normative change in raw score per year of age, and the standard-score coefficient gives the change in raw score per standard-score point. Their ratio gives the number of years of normative aging equivalent to one standard-score point. A woman’s cognitive age gap is that ratio multiplied by her own departure from the standard-score mean of 100, and her cognitive age is her chronological age plus that gap. A gap of zero denotes performance exactly as expected for her age, a positive gap performance typical of an older woman, and a negative gap that of a younger one. As speed measures decline more steeply with age than memory measures, a given standardized decrement converts to more years on speed domains, and the years metric is therefore reported throughout alongside the underlying standardized effect.

Importantly, cognitive age is an interpretive rescaling of the observed cognitive-performance difference as opposed to an independent outcome. All inferences regarding significantly poorer cognition are therefore evident in the original age-adjusted standard scores and do not depend on the age-equivalent transformation. Cognitive age is also meaningful only within the age range the normative data describe. A woman scoring far below the mean would otherwise be assigned a cognitive age below zero, and one scoring far above it an age beyond any human lifespan. These values would mean only that performance fell outside what the reference data can represent, and we therefore constrained every estimate to the 18 to 90 years range that the normative data cover. Since the bounds are defined relative to a woman’s own age, the lower bound binds younger women more tightly, and the uninjured band is the youngest in the cohort. Bounding therefore raises the mean in that band and lowers it in the most exposed one. Before bounding, mean gaps across the four bands were 7.4, 7.8, 18.7 and 24.4 years, and after bounding they are 9.5, 8.7, 17.3 and 20.8 years. Bounding compresses the gradient, so the reported estimates are conservative. Separately, derived age metrics can carry a regression artifact whereby the estimated gap correlates with chronological age, as described for brain-age estimates [87]. We tested for this directly and found none: the cognitive age gap was not associated with chronological age (*r* = −0.08, *P* = 0.27), as expected when scores are age-adjusted before conversion.

In addition, domains differ in how steeply raw performance declines with age, so the same standard-score deficit converts to more years in some domains than others. Certain cognitive skills, such as simple attention, are largely preserved across the lifespan and are not expected to change much as one ages. The slope relating raw performance to age in these domains is therefore relatively flat, and because the conversion divides by that slope, a given deficit translates into an exceedingly large number of years. Large deficits on these cognitive domains consequently yield substantial cognitive age gaps. We therefore interpret the larger estimates as indicating that performance falls outside the range the normative data describe, rather than as precise age equivalences. As noted above, we bound these estimates to the 18-to-90 year range the normative data cover, which truncates the largest gaps from above and therefore makes the reported values conservative. However, we acknowledge that it remains difficult to equate a severe impairment of an otherwise age-stable ability like simple attention to a numerical age, and the domains whose performance declines steadily with age, such as psychomotor speed and processing speed, therefore yield the most interpretable conversions.

##### Exposure bands

Participants were classified as reporting no partner-inflicted brain injury (BI), 1–3 injuries, 4–9 injuries, or ≥10 injuries. These cut-points were selected for clinical interpretability and to ensure adequate precision within each group, with at least 23 participants in every band. To evaluate whether the findings depended on this particular categorization, we examined all 390 alternative four-band schemes formed from candidate cut-points between 1 and 25 injuries that placed at least 15 participants in every band. The cognitive-age gradient was statistically distinguishable from zero in 96% of these schemes, with *P* values ranging from .001 to .072 (median, .011). The prespecified scheme ranked 205th of 390 by effect size, placing it in the weaker half of the candidate schemes and confirming that it did not maximise the observed association. We further conducted a permutation test in which the best-fitting banding scheme was reselected within each of 5,000 permutations. This analysis accounted for the full set of alternative cut-points and continued to support the cognitive-age gradient (*P* = .006). Although several alternative schemes produced marginally larger effect estimates, they created narrower middle-exposure groups and less stable, non-monotonic estimates. Finally, abandoning bands altogether, we re-estimated the contrast at each of twelve injury thresholds. Effect sizes rose from d = 0.19 at one or more injuries to d = 0.63 at twelve or more, and eleven of the twelve thresholds yielded a difference distinguishable from zero (Table S4).

#### Covariates

Each model adjusted for the same eleven covariates selected for prior association with both injury exposure and cognitive performance: age, education, race, BIs sustained outside the relationship in adulthood, BIs sustained in childhood, post-traumatic stress (PCL-5) [88], depression (PHQ-9) [89], anxiety (GAD-7) [90], alcohol use (AUDIT) [91], childhood maltreatment (Childhood Trauma Questionnaire [92], and abuse severity over the past twelve months (Composite Abuse Scale) [93,94]. Abuse severity was indexed by the frequency of abusive behaviors over the past twelve months, the interval the Composite Abuse Scale is designed to assess: the concern this covariate addresses is that ongoing abuse rather than past injury depresses test performance, so the relevant window is the one closest to assessment. Childhood injury includes violent shaking, consistent with the treatment of partner-inflicted shaking, so that the same definition applies on both sides of the model. Non-partner and childhood injury counts were log-transformed and defined identically to the primary exposure, including violent shaking.

Individually, these measures showed weak associations with cognitive measures, including psychomotor speed: childhood maltreatment (*r* = −0.17, *P* = 0.017), post-traumatic stress (*r* = −0.15, *P* = 0.041), depression (*r* = −0.14, *P* = 0.051), abuse severity (*r* = −0.13, *P* = 0.060), anxiety (*r* = −0.12, *P* = 0.101), and alcohol use (*r* = −0.04, *P* = 0.594). None was of a magnitude that could account for the association with injury count, which was *r* = −0.24 before any of them was entered and −0.22 with all of them adjusted (Table S8). In addition, only 10 women were in a current abusive relationship at assessment, and adjusting for a current abusive relationship did not affect the association, so the cognitive deficit does not appear to reflect ongoing threat.

#### Reporting

This observational study is reported in accordance with the STROBE statement for cross-sectional studies [95]. A completed STROBE checklist is provided with the submission. The study was approved by the institutional review board of Mass General Brigham, and all participants gave informed consent.

#### Statistical analysis

The study addressed whether the number of partner-inflicted brain injuries a woman sustained predicts her cognitive performance. The primary outcome was cognitive performance across the sixteen domains the battery reports, specified as a set rather than as a single index because the question concerns cognition broadly and no one domain was expected *a priori* to carry the association. We report each association with its confidence interval, and all tests were two-sided at an alpha of 0.05.

The primary exposure was the natural log of injury count, winsorized at 25. For each of the sixteen cognitive domains, we fitted a linear regression with the domain score as the outcome and injury count and the eleven covariates as predictors. The coefficient on injury count was tested by Wald test and is reported as a partial correlation, so that effects are comparable across domains. Six of the sixteen reached *P* < 0.05, against 0.8 expected were no association present.

Under Benjamini-Hochberg control across the sixteen domains, four associations remain significant at *q* < 0.05 (psychomotor speed, reaction time, processing speed and motor speed). Cognitive age was derived separately for each domain that reports a raw score, and is presented for those showing a significant association with injury count. Analyses use complete cases, and the number of observations contributing to each estimate is reported with that estimate. All remaining analyses are secondary or sensitivity analyses.

The sixteen domain associations are reported as effect estimates with confidence intervals and unadjusted *P* values, and interpreted as one profile rather than as sixteen separate tests; six reached P < 0.05 against 0.8 expected were no association present, with no domain treated as established or excluded on the basis of a threshold alone [96]. Comparisons of domain means against population norms were corrected by Bonferroni across the sixteen domains. Secondary analyses, including the comparison repeated at each cut-off from one to twelve injuries, alternative handling of extreme counts, alternative covariate sets, and restriction to mild BIs only, are reported as effect estimates with confidence intervals and unadjusted *P* values. These secondary analyses re-estimate a single contrast under alternative analytic choices rather than testing new hypotheses.

Brain injury count was severely right-skewed (median 1, interquartile range 0 to 4, range 0 to 1,095; adjusted Fisher-Pearson *G1* = 11.0) and is therefore summarized by median and interquartile range and modeled as log(1 + count) after capping at 25. Among plausible exposure codings, every alternative except the binary any-injury contrast fitted the data comparably by Akaike information criterion, the logarithmic specification falling within 0.24 units of the best-fitting coding (Fig. S1). As the functional form was selected empirically, we tested whether the inference depended on it using prespecified exposure bands, uncapped and untransformed counts, ordinal injury-frequency ratings, and rank-based analyses; the association was consistent across specifications, and the uncapped and untransformed alternatives are reported in Table S5.

Cognitive standard scores were approximately normally distributed (global cognition, Shapiro-Wilk *W* = 0.98) and are summarized by mean and standard deviation, in standard-score units with a normative mean of 100 and standard deviation of 15. Cognitive age gaps are reported in years. Group comparisons used Welch unpaired *t* tests, which do not assume equal variances, with Cohen’s d and 95% confidence intervals. Cohen’s *d* is reported unsigned throughout, with the direction of each difference given by the group means. Comparisons against population norms used one-sample *t*-tests against a hypothetical mean of 100. Effect sizes are referenced to the distribution each comparison is made against. For comparisons with population norms, *d* is the deviation of the group mean from 100 divided by the normative standard deviation of 15, so that it reads directly as a position in the normative distribution. For comparisons between groups within the cohort, *d* is the difference in means divided by the pooled standard deviation of the two groups compared. Sample sizes and t statistics are reported for every domain and every group in Tables S2 and S3 for improved replicability.

Trend across ordered exposure bands was tested by linear contrast. Bootstrap procedures used 4,000 to 10,000 resamples with a fixed seed. No data were imputed. Analyses use complete cases, and the number of observations contributing to each estimate is stated with that estimate; coverage ranges from 169 of 207 for global cognition, which requires every component subtest to be valid, to 206 for reasoning, and 206 of 207 women are complete on all eleven covariates. Sources of missing cognitive data, and the consequences of the exclusion rule that generates most of it, are examined directly in the Results and in Fig. 5.

Analyses were conducted in Python 3.12 and GraphPad Prism 11.0.2. The deposited dataset is provided as Data S1. Sample size was fixed by the size of the cohort and was not determined by power calculation. With 63 uninjured women and 32 reporting ten or more injuries, the smallest within-cohort difference detectable at 80% power is *d* = 0.61. Where a comparison between injury groups on a given cognitive domain does not reach significance, we describe it as no difference detected rather than as no difference, and we report confidence intervals so that readers can see how large a difference remains possible.

### Supplementary Text

#### Robustness to analytic choices

We tested whether the association was robust to alternative analytic decisions and potential sources of bias, including the influence of individual participants and adjustment for additional covariates.

Our primary analysis winsorized partner-inflicted BI counts at 25, following prior work in this cohort [42], as well as log-transformed to address extreme right-skewness (adjusted Fisher-Pearson *g₁* = 11.0). This was done to prevent extreme counts from disproportionately influencing continuous analyses. Consequently, fifteen women who reported more than 25 injuries, with counts reaching 1,095, all were assigned a value of 25 in the primary continuous analysis. To test whether this decision to cap the exposure affected the association, we repeated the analysis using the full reported counts. The association was slightly stronger when counts were left uncapped (partial *r* = −0.24, *P* = 0.0009). As a separate robustness analysis, we then excluded the fifteen women who exceeded the cap entirely. The association again remained essentially unchanged (Table S5).

Relatedly, to test whether the association was driven by a small number of influential participants, we examined Cook’s distance and standardised DFBETAs for the group coefficient in a model of cognitive age gap on ten-or-more versus no injury. No single participant exerted significant influence (maximum Cook’s D = 0.083; maximum absolute standardised DFBETA = 0.331). The corresponding values for the exposure coefficient of the primary adjusted model were 0.067 and 0.358.

Our primary analysis adjusted for eleven covariates. Two further covariates were tested in the subsample for whom they were recorded. Adding both covariates minimally strengthened the association (partial *r* = −0.27, *P* = 0.071 when excluded; partial *r* = −0.30, *P* = 0.046 when included). Neither neurological disorder (*r* = −0.001) nor current cannabis use (*r* = 0.073) was associated with injury count, so neither can account for the result. These covariates were not added to the primary model because they are recorded for less than a third of the sample, and doing so would have cost the majority of the observations to adjust for variables that do not confound the exposure.

Finally, our primary analysis grouped women into none, one to three, four to nine, and ten or more partner-inflicted BIs. To test whether the observed dose-response relationship between injury burden and cognitive performance depended on these cut-points, we repeated the analysis using four alternative grouping schemes (e.g., one to two rather than one to three; seven or more rather than ten or more as the highest-exposure band), varying the lower and upper boundaries and, in one analysis, dividing exposure into five bands. Across all four specifications, the gradient remained similar between injury exposure bands and cognitive age gaps (*P* for trend = 0.005 to 0.022). Treating injury count continuously rather than grouping it likewise preserved the association (Spearman *ρ* = 0.14, *P* = 0.043).

#### Mechanism of injury

Splitting the cohort by mechanism, psychomotor speed was 93.5 in women with blunt-force injury only, 95.1 in women with strangulation-related injury only, and 88.8 in women with both, against 95.8 in women with neither. Women with blunt-force versus strangulation-related injury alone did not differ from one another (*d* = 0.08, *P* = 0.69), and no other cognitive domain distinguished these two groups. When blunt-force and strangulation injury counts were entered simultaneously, greater blunt-force injury burden remained associated with poorer psychomotor speed (partial *r* = –0.20, *P* = 0.008), whereas strangulation-related injury burden did not (partial *r* = −0.05, *P* = 0.52). Similarly, psychomotor speed declined across increasing bands of blunt-force injury count (*P* for trend = 0.018), but not across bands of strangulation-related injury count (*P* = 0.25). These comparisons were substantially better powered for blunt-force injury, however, because only 16 women had strangulation-related injury without any blunt-force injury, so the failure to resolve a strangulation effect here should not be read as evidence that strangulation is cognitively harmless. Notably, women exposed to both mechanisms performed worst, suggesting that cumulative injury burden may be more important than injury mechanism alone.

### Supplementary Figures

**Fig. S1.**
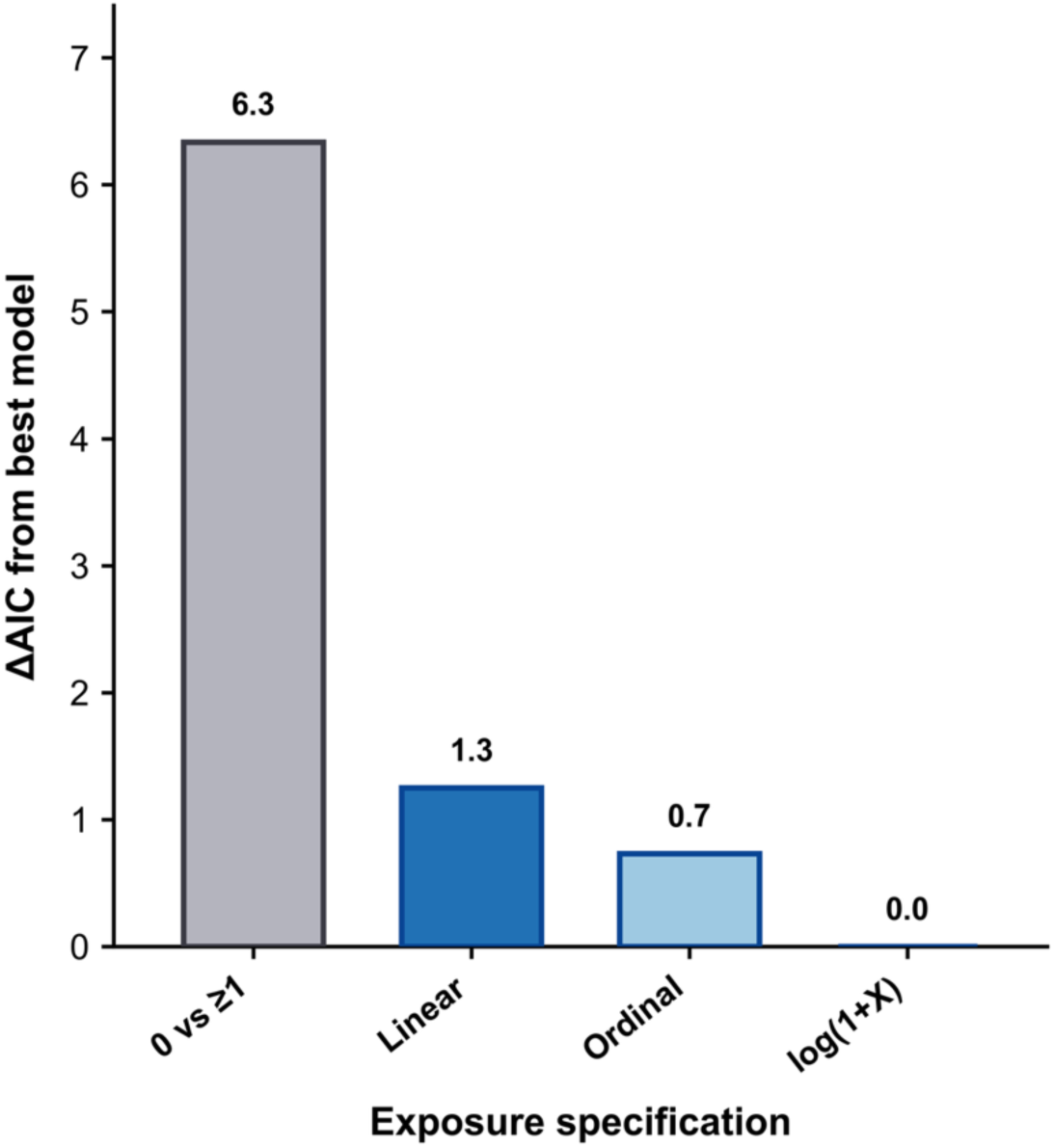
Model fit under alternative codings of injury exposure. Bars give the difference in Akaike information criterion (AIC) between each coding of partner-inflicted injury and the best-fitting coding, in age-adjusted models of psychomotor speed (*n* = 197). Lower AIC indicates better fit. Injury counts are capped at 25, as in the primary analysis. The logarithmic coding fits best (AIC = 1144.1), followed closely by the ordinal bands (1144.8) and the linear count (1145.3). Notably, the binary any-injury specification, the approach most often used in this literature, fits worst (1150.4). Leaving the counts uncapped does not change this. The logarithmic coding was therefore used, and the sensitivity analyses in Tables S4 to S7 show the association does not depend on this choice.

**Fig. S2.**
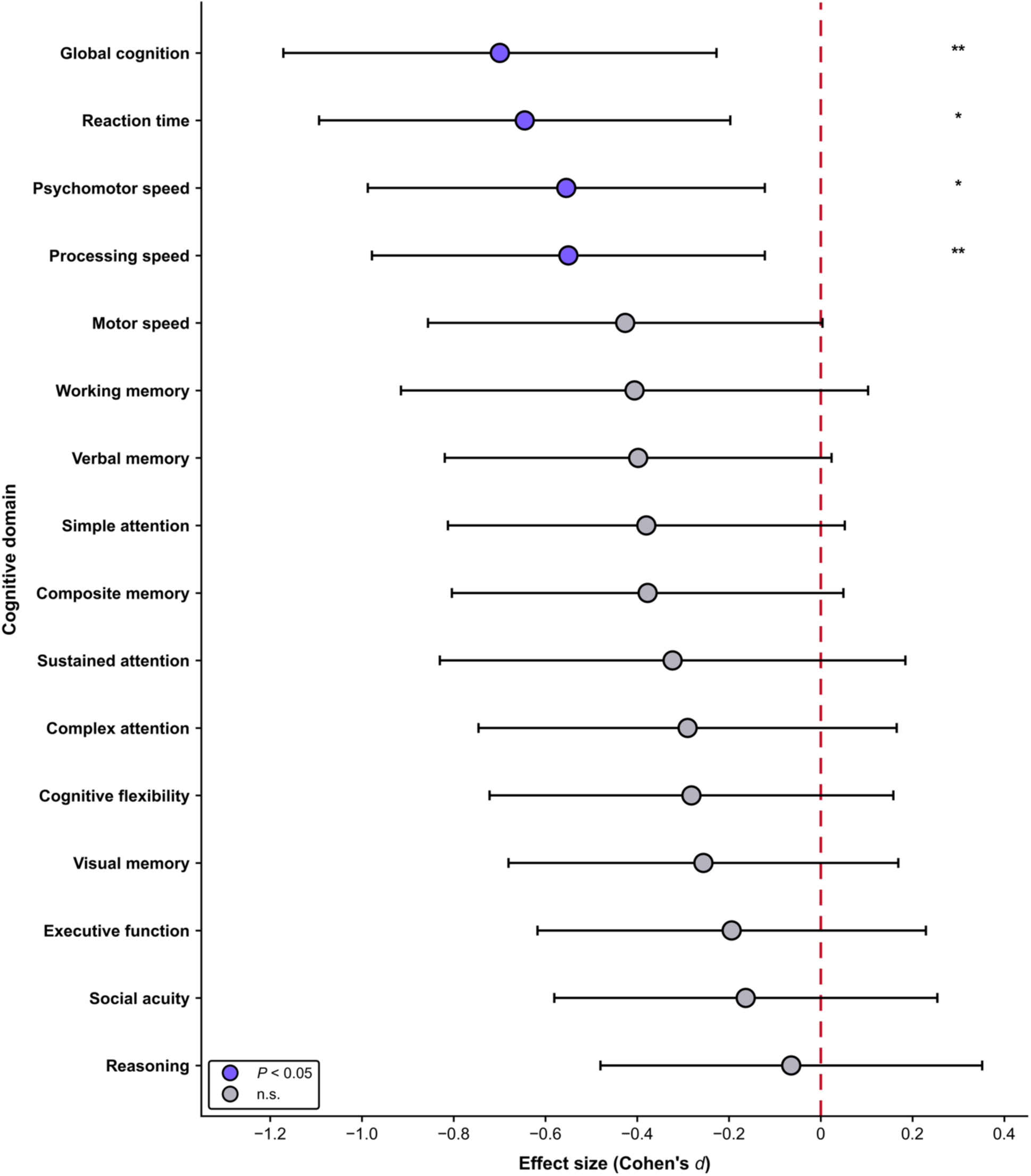
All sixteen cognitive domains ranked by effect size. Standardized difference between women reporting ten or more partner-inflicted brain injuries and women reporting none, with 95% confidence intervals. Global cognition shows the largest difference, followed by reaction time, psychomotor speed and processing speed (*P <* 0.05). The remaining twelve cognitive domains do not.

### Supplementary Tables

**Table S1.** Sample characteristics. Values are mean (SD) unless stated. *P* values compare women reporting ten or more partner-inflicted brain injuries against women reporting none, by Welch unpaired t-test. Group sizes in the header apply to the demographic, injury and psychiatric rows; each cognition row rests on a smaller complete-case sample, reported per domain in Table S3. Standard scores have a population mean of 100 and standard deviation of 15.

| Characteristic | All (*n* = 207) | No injury (*n* = 68) | 10 or more (*n* = 34) | *P* |
| --- | --- | --- | --- | --- |
| **Demographics** |  |  |  |  |
| Age, years | 41.0 (13.2) | 38.9 (14.5) | 40.6 (11.2) | 0.532 |
| Education, years | 15.7 (2.8) | 16.7 (2.6) | 15.6 (3.1) | 0.071 |
| Non-White, % | 30 | 28 | 29 | 1.000 |
| Hispanic, % | 16 | 19 | 18 | 1.000 |
| **Partner-inflicted brain injury** |  |  |  |  |
| Any injury, % | 67 | 0 | 100 |  |
| Repetitive injury, % | 42 | 0 | 100 |  |
| Strangulation-related, % | 27 | 0 | 53 |  |
| Exceeding the 25-event cap, % | 7 | 0 | 44 |  |
| Injuries, median (IQR) | 1 (0 to 4) | 0 (0 to 0) | 22 (12 to 40) |  |
| **Psychiatric and abuse measures** |  |  |  |  |
| Post-traumatic stress (PCL-5) | 33.5 (19.3) | 30.2 (18.0) | 41.9 (16.4) | 0.002 |
| Depression (PHQ-9) | 9.6 (5.8) | 8.5 (5.7) | 11.0 (5.2) | 0.033 |
| Anxiety (GAD-7) | 8.3 (5.7) | 7.7 (5.2) | 10.4 (5.6) | 0.019 |
| Alcohol use (AUDIT) | 3.4 (5.2) | 2.8 (3.3) | 3.3 (4.8) | 0.606 |
| Childhood maltreatment (CTQ) | 58.3 (22.1) | 53.8 (21.8) | 64.9 (20.0) | 0.013 |
| Abuse severity (CAS) | 6.9 (11.1) | 6.5 (10.9) | 10.6 (15.1) | 0.165 |
| Currently with an abusive partner (*n*) | 10 | 4 | 2 |  |
| **Cognition, standard score** |  |  |  |  |
| Global cognition | 93.8 (11.4) | 95.0 (9.7) | 87.6 (11.7) | 0.007 |
| Psychomotor speed | 93.5 (18.3) | 95.8 (17.0) | 86.2 (17.9) | 0.015 |
| Reaction time | 88.9 (18.0) | 90.9 (14.9) | 79.6 (21.8) | 0.014 |
| Processing speed | 97.7 (15.6) | 100.3 (17.7) | 91.2 (13.7) | 0.006 |
| Motor speed | 93.2 (17.6) | 94.8 (14.9) | 88.0 (17.9) | 0.070 |
| Composite memory | 92.1 (18.4) | 93.9 (17.6) | 86.9 (20.7) | 0.104 |
| Verbal memory | 92.6 (18.4) | 95.0 (18.3) | 87.8 (17.8) | 0.064 |
| Visual memory | 94.6 (16.4) | 95.3 (15.7) | 90.8 (20.4) | 0.284 |
| Complex attention | 96.9 (17.1) | 97.0 (16.0) | 91.9 (20.2) | 0.253 |
| Cognitive flexibility | 92.0 (16.6) | 91.7 (15.2) | 87.3 (16.4) | 0.224 |
| Executive function | 91.7 (16.7) | 90.3 (16.4) | 87.2 (16.0) | 0.368 |
| Social acuity | 99.8 (16.5) | 101.2 (15.4) | 98.8 (12.5) | 0.412 |
| Reasoning | 95.1 (16.0) | 94.9 (15.6) | 93.9 (15.6) | 0.763 |
| Working memory | 102.9 (11.8) | 105.0 (11.6) | 100.4 (10.8) | 0.113 |
| Sustained attention | 104.2 (9.5) | 105.5 (8.1) | 102.8 (9.4) | 0.245 |
| Simple attention | 90.9 (21.7) | 93.7 (20.1) | 85.7 (22.7) | 0.102 |
| Lost $\geq 1$ score to validity criteria, % | 40 | 43 | 38 | — |

**Table S2.** All sixteen cognitive domains against population norms across whole cohort and among those with history of partner-inflicted brain injury. One-sample *t* tests against a normative mean of 100, with Bonferroni correction across sixteen tests. Sample sizes vary across domains because scores flagged as invalid by the battery’s validity indicator were excluded, leaving 143 to 206 of the 207 women per domain. Global cognition additionally requires every component subtest to be valid, therefore resulting in a lower sample size.

|  | Whole cohort |  |  |  | ≥ 1 partner-inflicted BI |  |  |  | ≥ 10 partner-inflicted BIs |  |  |  |
| --- | --- | --- | --- | --- | --- | --- | --- | --- | --- | --- | --- | --- |
| Domain | n | Mean | t | P<sub>Bonferroni</sub> | n | Mean | t | P<sub>Bonferroni</sub> | n | Mean | t | P<sub>Bonferroni</sub> |
| Reaction time | 190 | 88.9 | -8.48 | <0.0001 | 130 | 88.0 | -7.10 | <0.0001 | 30 | 79.6 | -5.12 | 0.0003 |
| Simple attention | 192 | 90.9 | -5.82 | <0.0001 | 128 | 89.5 | -5.31 | <0.0001 | 31 | 85.7 | -3.51 | 0.0232 |
| Executive function | 199 | 91.7 | -7.03 | <0.0001 | 133 | 92.4 | -5.23 | <0.0001 | 32 | 87.2 | -4.53 | 0.0013 |
| Cognitive flexibility | 188 | 92.0 | -6.62 | <0.0001 | 128 | 92.1 | -5.15 | <0.0001 | 30 | 87.3 | -4.25 | 0.0032 |
| Composite memory | 201 | 92.1 | -6.11 | <0.0001 | 136 | 91.2 | -5.48 | <0.0001 | 32 | 86.9 | -3.59 | 0.0182 |
| Verbal memory | 204 | 92.6 | -5.71 | <0.0001 | 138 | 91.5 | -5.44 | <0.0001 | 33 | 87.8 | -3.92 | 0.0069 |
| Motor speed | 197 | 93.2 | -5.42 | <0.0001 | 134 | 92.5 | -4.65 | 0.0001 | 32 | 88.0 | -3.79 | 0.0103 |
| Psychomotor speed | 197 | 93.5 | -5.02 | <0.0001 | 134 | 92.4 | -4.71 | <0.0001 | 32 | 86.2 | -4.36 | 0.0021 |
| Global cognition | 169 | 93.8 | -7.10 | <0.0001 | 117 | 93.2 | -6.05 | <0.0001 | 28 | 87.6 | -5.58 | 0.0001 |
| Visual memory | 201 | 94.6 | -4.72 | <0.0001 | 136 | 94.2 | -4.04 | 0.0014 | 32 | 90.8 | -2.54 | 0.2585 |
| Reasoning | 206 | 95.1 | -4.39 | 0.0003 | 138 | 95.2 | -3.48 | 0.0109 | 33 | 93.9 | -2.24 | 0.5179 |
| Complex attention | 174 | 96.9 | -2.41 | 0.2690 | 118 | 96.8 | -1.95 | 0.8560 | 28 | 91.9 | -2.12 | 0.6945 |
| Processing speed | 204 | 97.7 | -2.08 | 0.6198 | 137 | 96.5 | -2.88 | 0.0747 | 32 | 91.2 | -3.63 | 0.0162 |
| Social acuity | 201 | 99.8 | -0.14 | 1.0000 | 134 | 99.1 | -0.58 | 1.0000 | 33 | 98.8 | -0.53 | 1.0000 |
| Working memory | 143 | 102.9 | 2.96 | 0.0584 | 95 | 101.9 | 1.55 | 1.0000 | 22 | 100.4 | 0.16 | 1.0000 |
| Sustained attention | 143 | 104.2 | 5.31 | <0.0001 | 95 | 103.5 | 3.42 | 0.0147 | 22 | 102.8 | 1.38 | 1.0000 |

**Table S3.** Standardized differences across all sixteen cognitive domains for women reporting ten or more partner-inflicted brain injuries versus women reporting none. *P* values are unadjusted.

| Domain | n (no /10+) | No injury | Ten or more | *d* | 95% CI | *P* |
| --- | --- | --- | --- | --- | --- | --- |
| Global cognition | 52/28 | 95.0 | 87.6 | 0.70 | 0.23, 1.17 | 0.007 |
| Reaction time | 60/30 | 90.9 | 79.6 | 0.64 | 0.20, 1.09 | 0.014 |
| Psychomotor speed | 63/32 | 95.8 | 86.2 | 0.55 | 0.12, 0.99 | 0.015 |
| Processing speed | 67/32 | 100.3 | 91.2 | 0.55 | 0.12, 0.98 | 0.006 |
| Motor speed | 63/32 | 94.8 | 88.0 | 0.43 | 0.00, 0.86 | 0.070 |
| Working memory | 48/22 | 105.0 | 100.4 | 0.41 | -0.10, 0.91 | 0.113 |
| Verbal memory | 66/33 | 95.0 | 87.8 | 0.40 | -0.02, 0.82 | 0.064 |
| Composite memory | 65/32 | 93.9 | 86.9 | 0.38 | -0.05, 0.80 | 0.104 |
| Simple attention | 64/31 | 93.7 | 85.7 | 0.38 | -0.05, 0.81 | 0.102 |
| Sustained attention | 48/22 | 105.5 | 102.8 | 0.32 | -0.18, 0.83 | 0.245 |
| Complex attention | 56/28 | 97.0 | 91.9 | 0.29 | -0.17, 0.75 | 0.253 |

| Domain | *n* (no /10+) | No injury | Ten or more | *d* | 95% CI | *P* |
| --- | --- | --- | --- | --- | --- | --- |
| Cognitive flexibility | 60/30 | 91.7 | 87.3 | 0.28 | -0.16, 0.72 | 0.224 |
| Visual memory | 65/32 | 95.3 | 90.8 | 0.26 | -0.17, 0.68 | 0.284 |
| Executive function | 66/32 | 90.3 | 87.2 | 0.19 | -0.23, 0.62 | 0.368 |
| Social acuity | 67/33 | 101.2 | 98.8 | 0.16 | -0.25, 0.58 | 0.412 |
| Reasoning | 68/33 | 94.9 | 93.9 | 0.06 | -0.35, 0.48 | 0.763 |

**Table S4.** Sensitivity of the effect to the injury count used to define high exposure. Twelve thresholds were tested, with psychomotor speed as the outcome. Each threshold is compared against women reporting no partner-inflicted brain injury.

| Threshold defining high exposure | *n exposed* | *d* | 95% CI | *P* |
| --- | --- | --- | --- | --- |
| 1 or more | 134 | 0.19 | -0.11, 0.49 | 0.200 |
| 2 or more | 83 | 0.36 | 0.03, 0.69 | 0.032 |
| 3 or more | 65 | 0.39 | 0.04, 0.74 | 0.028 |
| 4 or more | 54 | 0.46 | 0.09, 0.83 | 0.015 |
| 5 or more | 45 | 0.61 | 0.22, 1.00 | 0.003 |
| 6 or more | 40 | 0.53 | 0.13, 0.94 | 0.012 |
| 7 or more | 39 | 0.53 | 0.13, 0.94 | 0.012 |
| 8 or more | 37 | 0.50 | 0.09, 0.91 | 0.022 |
| 9 or more | 34 | 0.48 | 0.06, 0.90 | 0.032 |
| 10 or more | 32 | 0.55 | 0.12, 0.99 | 0.015 |
| 11 or more | 28 | 0.57 | 0.12, 1.03 | 0.019 |
| 12 or more | 26 | 0.63 | 0.16, 1.10 | 0.014 |

**Table S5.** Sensitivity to the handling of extreme injury counts. The primary analysis caps counts at 25; alternatives exclude women above the stated value. Gap is the mean cognitive age gap for women reporting ten or more injuries against the general population, bounded to ages 18 to 90 as in the main analyses; d is the standardized difference in psychomotor speed against women reporting none. Every alternative preserves the direction of the effect, with gaps ranging from 19.0 to 24.6 years and d from 0.45 to 0.67 against 0.55 in the primary analysis.

| Handling of extreme counts | *n* | Gap (years) | *d* | 95% CI |
| --- | --- | --- | --- | --- |
| Capped at 25 (primary) | 207 | 20.8 | 0.55 | 0.12 to 0.99 |
| Exclude counts above 145 | 202 | 19.1 | 0.45 | 0.00 to 0.91 |
| Exclude counts above 100 | 202 | 19.1 | 0.45 | 0.00 to 0.91 |
| Exclude counts above 50 | 201 | 19.0 | 0.45 | -0.01 to 0.91 |
| Exclude counts above 25 | 192 | 24.6 | 0.67 | 0.13 to 1.22 |
| Exclude counts above 16 | 188 | 23.3 | 0.64 | 0.05 to 1.22 |

**Table S6.** Sensitivity to the covariate set. Partial correlations between log injury count and psychomotor speed under progressively fuller adjustment.

| Covariate set | *n* | Partial *r* | *P* |
| --- | --- | --- | --- |
| + Age | 197 | -0.198 | 0.0055 |
| + Education | 197 | -0.184 | 0.0099 |
| + Race | 197 | -0.182 | 0.0112 |
| + Non-partner adult brain injury | 197 | -0.240 | 0.0008 |
| + Childhood brain injury | 197 | -0.231 | 0.0012 |
| + Post-traumatic stress | 197 | -0.215 | 0.0028 |

| Covariate set | *n* | *Partial r* | *P* |
| --- | --- | --- | --- |
| + Depression | 197 | -0.222 | 0.0021 |
| + Anxiety | 197 | -0.223 | 0.0020 |
| + Alcohol use | 196 | -0.232 | 0.0014 |
| + Childhood maltreatment | 196 | -0.231 | 0.0015 |
| + Abuse severity | 196 | -0.221 | 0.0025 |

**Table S7.** Sensitivity to the injury definition. The primary definition counts all qualifying partner-inflicted brain injuries, including moderate-to-severe events, whereas the alternative definition counts only mild injuries. Under the mild-only definition, women whose injuries were exclusively moderate-to-severe are therefore classified as having no qualifying injury. Moderate-to-severe events are rare in this cohort, with nine events among eight women, and only one woman is reclassified from the exposed group to the reference group. That woman contributes no valid cognitive score, so every estimate below is identical under the two definitions, and this sensitivity analysis has little power to distinguish them. Gaps are bounded to ages 18 to 90 as in the primary analyses.

| Injury definition | *n* (0 / 10+) | Absolute gap | vs uninjured | *d* |
| --- | --- | --- | --- | --- |
| Total brain injury (primary) | 63/32 | 20.8 | 11.3 | 0.55 |
| Mild brain injury only | 63/32 | 20.8 | 11.3 | 0.55 |

**Table S8.** Alternative explanations tested directly. Each measure is related both to cognition and to injury count, adjusted for age. A confounder would have to predict both. Childhood maltreatment and post-traumatic stress are each weakly associated with cognition, but neither is of a magnitude that could account for the association with injury count.

| Measure | *n* | *r* with cognition | *P* | *r* with injury count | *P* |
| --- | --- | --- | --- | --- | --- |
| Post-traumatic stress | 197 | -0.146 | 0.041 | 0.213 | 0.003 |
| Depression | 197 | -0.140 | 0.051 | 0.121 | 0.090 |
| Anxiety | 197 | -0.120 | 0.093 | 0.149 | 0.037 |
| Alcohol use | 196 | -0.040 | 0.583 | 0.048 | 0.502 |
| Childhood maltreatment | 197 | -0.171 | 0.017 | 0.134 | 0.061 |
| Abuse severity | 197 | -0.135 | 0.059 | 0.172 | 0.016 |

**Table S9.** Cognitive domains and the subtests from which each is scored. Descriptions are provided verbatim from the CNS Vital Signs score report.

| Cognitive test | Derived CNSVS domain | CNSVS test description |
| --- | --- | --- |
| Verbal Memory Test (VBM) | Composite Memory | Fifteen target words are presented and must then be picked out from among fifteen distractors, with the recognition trial repeated at the end of the battery. The score is the number of target words correctly identified, indexing recognition and retrieval of verbal material; lower scores indicate weaker verbal memory. |
|  | Verbal Memory |  |
| Visual Memory Test (VSM) | Composite Memory | Fifteen geometric figures are presented and must then be picked out from among fifteen distractors, with the recognition trial repeated at the end of the battery. The score is the number of target figures correctly identified, indexing recognition and retrieval of visual material; lower scores indicate weaker visual memory. |
|  | Visual Memory |  |

| **Cognitive test** | **Derived CNSVS domain** | **CNSVS test description** |
| --- | --- | --- |
| **Finger Tapping Test (FTT)** | Psychomotor Speed | Three timed tapping trials are completed with each hand. The score is the number of taps per hand, indexing fine motor speed and control; fewer taps indicate motor slowing, and rates differ systematically between the dominant and non-dominant hand. |
|  | Motor Speed |  |
| **Symbol Digit Coding (SDC)** | Processing Speed | Digits are matched to symbols against the clock. Performance depends jointly on visual scanning, perception, short-term visual memory and motor output, so the score indexes processing speed broadly rather than any single component; errors may arise from impulsive responding as well as from misperception. |
|  | Psychomotor Speed |  |
| **Stroop Test (ST)** | Reaction Time | Simple and complex reaction times are recorded across congruent and incongruent colour-word conditions, indexing response inhibition and the capacity to keep pace as the instructions change and grow more demanding. Longer latencies indicate slowed processing. |
|  | Complex Attention |  |
| **Shifting Attention Test (SAT)** | Cognitive Flexibility | Matching rules change without warning, so responses must be re-mapped while competing rules are held in mind. The score combines correct responses, errors and response latency, and indexes set-shifting and the handling of concurrent task demands. |
|  | Executive Function |  |
|  | Complex Attention |  |
| **Continuous Performance Test (CPT)** | Simple Attention | Responses are made to a designated target over a sustained interval, yielding measures of vigilance and choice reaction time. Unimpaired performance is close to ceiling, so a small number of errors, or unusually long latencies, carry diagnostic weight. |
|  | Complex Attention |  |
| **Perception of Emotions Test (POET)** | Social Acuity | Facial expressions are classified as happy, calm, angry or sad, indexing the perception and identification of emotion. Response latencies are considerably longer than on the other subtests, reflecting the additional processing that emotion recognition requires. |
| **Non-Verbal Reasoning Test (NVRT)** | Reasoning | Fifteen matrix problems of increasing difficulty are presented, each for 14.5 seconds, indexing the perception of visual-abstract information and the recognition of relationships among its elements. |
| **Four-Part Continuous Performance Test (FPCPT)** | Working Memory | Four parts of increasing load: a simple reaction-time block; a continuous-performance discrimination block yielding choice reaction times; a one-back block; and a two-back block. The two-back block yields the working memory score, and the second, third and fourth blocks together yield sustained attention. |
|  | Sustained Attention |  |

**Table S10.** Association between injury count and each speed and memory domain, adjusted for age. Speed domains vary with injury count more strongly than memory domains, the reverse of the pattern memory-driven under-reporting would produce. Averaged across domains, the age-adjusted association is larger for speed (0.19) than for memory (0.12). The two sets share participants, so we report the contrast descriptively rather than testing it.

| Domain type | Domain | *n* | \|*r*\| with injury count | *P* |
| --- | --- | --- | --- | --- |
| Speed | Psychomotor speed | 197 | 0.198 | 0.005 |
| Speed | Motor speed | 197 | 0.151 | 0.035 |
| Speed | Processing speed | 204 | 0.202 | 0.004 |
| Speed | Reaction time | 190 | 0.210 | 0.004 |
| Memory | Composite memory | 201 | 0.131 | 0.064 |
| Memory | Verbal memory | 204 | 0.138 | 0.050 |
| Memory | Visual memory | 201 | 0.095 | 0.183 |
|  | Speed mean |  | 0.190 |  |
|  | Memory mean |  | 0.121 |  |

**Table S11.** Cognitive association by injury mechanism. Partial correlations between psychomotor speed and the log count of each injury mechanism, adjusted for the eleven covariates of the primary model, entered first separately and then together. Total injury count is not entered, because blunt-force events constitute 89% of all injuries and the two are collinear (*r* = 0.998 for the raw counts, and 0.884 for the log-transformed predictors actually entered), so a model containing both is uninterpretable. Strangulation injuries were reported by 56 women with a median of two, so this comparison is better powered to detect an effect of blunt-force injury.

| Injury mechanism | *n* | *Partial r* | *P* |
| --- | --- | --- | --- |
| Blunt-force injuries, entered alone | 196 | −0.208 | 0.0046 |
| Strangulation injuries, entered alone | 196 | −0.086 | 0.2457 |
| Blunt-force injuries, adjusted for strangulation | 196 | −0.195 | 0.0079 |
| Strangulation injuries, adjusted for blunt force | 196 | −0.047 | 0.5220 |

**Table S12.** Cognitive age gap by domain. Cognitive age was derived separately for each domain by regressing the raw score on chronological age and the age-adjusted standard score, converting the standard-score deficit into years, and bounding the resulting cognitive age to 18 to 90 years. Values give the gap for women reporting ten or more partner-inflicted brain injuries, against the general population and against women in this cohort reporting none. The years-per-point column gives the conversion factor, which is the number of years of age equivalent to one standard-score point in that domain. Conversion factors vary widely because domains differ in how steeply their raw scores decline with age, and estimates from domains with shallow slopes, such as simple attention and social acuity, are correspondingly less precise. Two-sided t tests, unadjusted. Cognitive age was derived for each of the fifteen domains that report a raw score. Given that global cognition is an average of age-standardized scores across multiple cognitive domains, and therefore has no raw performance measure that changes with age, a cognitive age for global cognition cannot be calculated.

| Domain | Years per point | *n* | Gap vs population (95% CI) | *P* | Gap within cohort (95% CI) | *P* |
| --- | --- | --- | --- | --- | --- | --- |
| Simple attention | 32.79 | 31 | 28.1 (14.9 to 41.4) | 0.0002 | 14.2 (−1.1 to 29.5) | 0.074 |
| Reaction time | 2.04 | 30 | 27.8 (18.1 to 37.4) | <0.0001 | 11.9 (1.1 to 22.7) | 0.036 |
| Verbal memory | 5.31 | 33 | 26.0 (15.8 to 36.2) | <0.0001 | 13.7 (1.2 to 26.3) | 0.036 |
| Motor speed | 4.50 | 32 | 24.6 (13.6 to 35.6) | <0.0001 | 6.0 (−6.6 to 18.5) | 0.353 |
| Composite memory | 4.14 | 32 | 22.7 (12.4 to 33.0) | <0.0001 | 6.2 (−6.2 to 18.7) | 0.331 |
| Cognitive flexibility | 2.24 | 30 | 22.6 (13.0 to 32.1) | <0.0001 | 8.0 (−2.9 to 18.9) | 0.158 |
| Executive function | 2.19 | 32 | 21.9 (13.3 to 30.6) | <0.0001 | 6.3 (−3.8 to 16.5) | 0.227 |
| **Psychomotor speed** | 1.77 | 32 | 20.8 (11.6 to 29.9) | <0.0001 | 11.3 (1.2 to 21.5) | 0.033 |
| **Visual memory** | 4.57 | 32 | 19.7 (8.4 to 31.1) | 0.0013 | 1.7 (-11.7 to 15.1) | 0.807 |
| **Reasoning** | 5.65 | 33 | 15.9 (5.8 to 25.9) | 0.0030 | 1.7 (-10.1 to 13.5) | 0.783 |
| **Complex attention** | 4.41 | 28 | 11.4 (-2.5 to 25.3) | 0.1033 | 2.2 (-13.2 to 17.7) | 0.779 |
| **Processing speed** | 1.30 | 32 | 11.4 (5.0 to 17.8) | 0.0010 | 8.6 (1.3 to 15.8) | 0.024 |
| **Social acuity** | 25.46 | 33 | 11.2 (-3.2 to 25.6) | 0.1220 | 3.9 (-12.6 to 20.4) | 0.644 |
| **Working memory** | 6.31 | 22 | 5.0 (-7.8 to 17.8) | 0.4277 | 4.6 (-10.0 to 19.1) | 0.542 |
| **Sustained attention** | 6.45 | 22 | -6.8 (-17.3 to 3.6) | 0.1883 | -2.2 (-14.3 to 9.8) | 0.717 |

## References

1. H. Zhang Kudon, S. Zhu, B. Chen, M. J. Breiding, R. W. Leemis, X. Zhang, A. Schwank, K. C. Basile, The National Intimate Partner and Sexual Violence Survey (NISVS): 2023/2024 Intimate Partner Violence Data Brief (Centers for Disease Control and Prevention, National Center for Injury Prevention and Control, 2026); https://www.cdc.gov/nisvs/media/pdfs/intimatepartnerviolence-brief.pdf.

2. K. Le, M. Nguyen, War and Intimate Partner Violence in Africa. Sage Open 12, 21582440221096427 (2022).

3. S. Svallfors, Hidden Casualties: The Links between Armed Conflict and Intimate Partner Violence in Colombia. Politics & Gender 19, 133–165 (2023).

4. D. Schneider, K. Harknett, S. McLanahan, Intimate Partner Violence in the Great Recession. Demography 53, 471–505 (2016).

5. M. Kyriakidou, A. Zalaf, S. Christophorou, A. Ruiz-Garcia, C. Valanides, Longitudinal Fluctuations of National Help-Seeking Reports for Domestic Violence Before, During, and After the Financial Crisis in Cyprus. J Interpers Violence 36, NP8333–NP8346 (2021).

6. E. Arenas-Arroyo, D. Fernandez-Kranz, N. Nollenberger, Intimate partner violence under forced cohabitation and economic stress: Evidence from the COVID-19 pandemic. Journal of Public Economics 194, 104350 (2021).

7. A. R. Piquero, W. G. Jennings, E. Jemison, C. Kaukinen, F. M. Knaul, Domestic violence during the COVID-19 pandemic - Evidence from a systematic review and meta-analysis. Journal of Criminal Justice 74, 101806 (2021).

8. E. M. Allen, L. Munala, J. R. Henderson, Kenyan Women Bearing the Cost of Climate Change. International Journal of Environmental Research and Public Health 18, 12697 (2021).

9. Y. Guo, Y. Zhu, Z. Fatmi, L. Zhou, C. He, J. Bachwenkizi, H. Kan, R. Chen, Flood exposure and intimate partner violence in low- and middle-income countries. Nat Water 3, 296–306 (2025).

10. M. Anastario, N. Shehab, L. Lawry, Increased Gender-based Violence Among Women Internally Displaced in Mississippi 2 Years Post–Hurricane Katrina. Disaster Medicine and Public Health Preparedness 3, 18–26 (2009).

11. J. A. Schumacher, S. F. Coffey, F. H. Norris, M. Tracy, K. Clements, S. Galea, Intimate Partner Violence and Hurricane Katrina: Predictors and Associated Mental Health Outcomes. Violence and Victims 25, 588–603 (2010).

12. E. W. Harville, C. A. Taylor, H. Tesfai, X. Xiong, P. Buekens, Experience of Hurricane Katrina and Reported Intimate Partner Violence. J Interpers Violence 26, 833–845 (2011).

13. B. Sanz-Barbero, C. Linares, C. Vives-Cases, J. L. González, J. J. López-Ossorio, J. Díaz, Heat wave and the risk of intimate partner violence. Science of The Total Environment 644, 413–419 (2018).

14. Y. Zhu, C. He, M. Bell, Y. Zhang, Z. Fatmi, Y. Zhang, M. Zaid, J. Bachwenkizi, C. Liu, L. Zhou, R. Chen, H. Kan, Association of Ambient Temperature With the Prevalence of Intimate Partner Violence Among Partnered Women in Low- and Middle-Income South Asian Countries. JAMA Psychiatry 80, 952–961 (2023).

15. R. J. McCarthy, M. M. Rabenhorst, J. S. Milner, W. J. Travis, P. S. Collins, What difference does a day make? Examining temporal variations in partner maltreatment. J Fam Psychol 28, 421–428 (2014).

16. S. Kirby, B. Francis, R. O’Flaherty, Can the FIFA World Cup Football (Soccer) Tournament Be Associated with an Increase in Domestic Abuse? Journal of Research in Crime and Delinquency 51, 259–276 (2014).

17. B. Sanz-Barbero, C. Linares, C. Vives-Cases, J. L. González, J. J. López-Ossorio, J. Díaz, Intimate partner violence in Madrid: a time series analysis (2008–2016). Annals of Epidemiology 28, 635–640 (2018).

18. L. E. Kwako, N. Glass, J. Campbell, K. C. Melvin, T. Barr, J. M. Gill, Traumatic Brain Injury in Intimate Partner Violence: A Critical Review of Outcomes and Mechanisms. Trauma, Violence, & Abuse 12, 115–126 (2011).

19. H. (Lin) Haag, D. Jones, T. Joseph, A. Colantonio, Battered and Brain Injured: Traumatic Brain Injury Among Women Survivors of Intimate Partner Violence—A Scoping Review. Trauma, Violence, & Abuse 23, 1270–1287 (2022).

20. C. Esopenko, D. Jain, S. P. Adhikari, K. Dams-O’Connor, M. Ellis, H. (Lin) Haag, E. S. Hovenden, F. Keleher, I. K. Koerte, H. M. Lindsey, A. D. Marshall, K. Mason, J. S. McNally, D. S. Menefee, T. L. Merkley, E. N. Read, P. Rojcyk, S. R. Shultz, M. Sun, D. Toccalino, E. M. Valera, P. van Donkelaar, C. Wellington, E. A. Wilde, Intimate Partner Violence-Related Brain Injury: Unmasking and Addressing the Gaps. Journal of Neurotrauma 41, 2219–2237 (2024).

21. J. E. Karr, T. K. Logan, Post-Concussion Symptoms in Women With Head Injury Due to Intimate Partner Violence. Journal of Neurotrauma 41, 447–463 (2024).

22. G. M. Turner, C. McMullan, O. L. Aiyegbusi, D. Bem, T. Marshall, M. Calvert, J. Mant, A. Belli, Stroke risk following traumatic brain injury: Systematic review and meta-analysis. International Journal of Stroke 16, 370–384 (2021).

23. J. F. Annegers, W. A. Hauser, S. P. Coan, W. A. Rocca, A Population-Based Study of Seizures after Traumatic Brain Injuries. New England Journal of Medicine 338, 20–24 (1998).

24. C. Harrison-Felix, S. A. Kolakowsky-Hayner, F. M. Hammond, R. Wang, J. Englander, K. Dams-O’Connor, S. E. D. Kreider, T. A. Novack, R. Diaz-Arrastia, Mortality after surviving traumatic brain injury: risks based on age groups. J Head Trauma Rehabil 27, E45–56 (2012).

25. L. Wilson, L. Horton, K. Kunzmann, B. J. Sahakian, V. F. Newcombe, E. A. Stamatakis, N. von Steinbuechel, K. Cunitz, A. Covic, A. Maas, D. V. Praag, D. Menon, Understanding the relationship between cognitive performance and function in daily life after traumatic brain injury. J Neurol Neurosurg Psychiatry 92, 407–417 (2021).

26. A. I. R. Maas, D. K. Menon, G. T. Manley, M. Abrams, C. Åkerlund, N. Andelic, M. Aries, T. Bashford, M. J. Bell, Y. G. Bodien, B. L. Brett, A. Büki, R. M. Chesnut, G. Citerio, D. Clark, B. Clasby, D. J. Cooper, E. Czeiter, M. Czosnyka, K. Dams-O’Connor, V. D. Keyser, R. Diaz-Arrastia, A. Ercole, T. A. van Essen, É. Falvey, A. R. Ferguson, A. Figaji, M. Fitzgerald, B. Foreman, D. Gantner, G. Gao, J. Giacino, B. Gravesteijn, F. Guiza, D. Gupta, M. Gurnell, J. A. Haagsma, F. M. Hammond, G. Hawryluk, P. Hutchinson, M. van der Jagt, S. Jain, S. Jain, J. Jiang, H. Kent, A. Kolias, E. J. O. Kompanje, F. Lecky, H. F. Lingsma, M. Maegele, M. Majdan, A. Markowitz, M. McCrea, G. Meyfroidt, A. Mikolić, S. Mondello, P. Mukherjee, D. Nelson, L. D. Nelson, V. Newcombe, D. Okonkwo, M. Orešič, W. Peul, D. Pisică, S. Polinder, J. Ponsford, L. Puybasset, R. Raj, C. Robba, C. Røe, J. Rosand, P. Schueler, D. J. Sharp, P. Smielewski, M. B. Stein, N. von Steinbüchel, W. Stewart, E. W. Steyerberg, N. Stocchetti, N. Temkin, O. Tenovuo, A. Theadom, I. Thomas, A. T. Espin, A. F. Turgeon, A. Unterberg, D. V. Praag, E. van Veen, J. Verheyden, T. V. Vyvere, K. K. W. Wang, E. J. A. Wiegers, W. H. Williams, L. Wilson, S. R. Wisniewski, A. Younsi, J. K. Yue, E. L. Yuh, F. A. Zeiler, M. Zeldovich, R. Zemek, Traumatic brain injury: progress and challenges in prevention, clinical care, and research. The Lancet Neurology 21, 1004–1060 (2022).

27. E. M. Valera, I. Sanghvi, S. R. Sitto, J. Chua, A. Saadi, A. Theadom, “I Can Remember Thinking, Like Almost Wishing, That the Injuries Would Have Been Worse, Because Then I Wouldn’t Be Questioned”: A Qualitative Study on Women’s Experience of Accessing Healthcare for Intimate Partner Violence-Related Brain Injury. Healthcare 14, 165 (2026).

28. D. Toccalino, H. L. Haag, E. Nalder, V. Chan, A. Moore, A. Colantonio, C. M. Wickens, “A whole ball of all-togetherness”: The interwoven experiences of intimate partner violence, brain injury, and mental health. PLoS One 19, e0306599 (2024).

29. National Academies of Sciences, Engineering, and Medicine, Traumatic Brain Injury: A Roadmap for Accelerating Progress (National Academies Press, 2022).

30. M. A. Lindberg, E. M. Moy Martin, D. W. Marion, Military Traumatic Brain Injury: The History, Impact, and Future. J Neurotrauma 39, 1133–1145 (2022).

31. J. T. Parsons, C. M. Baugh, The evolving landscape of policies, rules, and law in sport-related concussion. Handb Clin Neurol 158, 257–267 (2018).

32. E. Hoch, J. Martinez, R. Bakhshi, M. Hieber, A. Presser, T. Tartaglia, A Review of U.S. Military Traumatic Brain Injury Studies: Trends, Gaps, and Opportunities. Rand Health Q 13, 7 (2026).

33. E. M. Valera, Increasing Our Understanding of an Overlooked Public Health Epidemic: Traumatic Brain Injuries in Women Subjected to Intimate Partner Violence. J Womens Health (Larchmt) 27, 735–736 (2018).

34. A. Saadi, L. Chibnik, E. Valera, Examining the Association Between Childhood Trauma, Brain Injury, and Neurobehavioral Symptoms Among Survivors of Intimate Partner Violence: A Cross-Sectional Analysis. J Head Trauma Rehabil 37, 24–33 (2022).

35. M. B. Stein, C. M. Kennedy, E. W. Twamley, Neuropsychological function in female victims of intimate partner violence with and without posttraumatic stress disorder. Biological Psychiatry 52, 1079–1088 (2002).

36. J. C. Campbell, J. C. Anderson, A. McFadgion, J. Gill, E. Zink, M. Patch, G. Callwood, D. Campbell, The Effects of Intimate Partner Violence and Probable Traumatic Brain Injury on Central Nervous System Symptoms. J Womens Health (Larchmt) 27, 761–767 (2018).

37. A. N. Cimino, G. Yi, M. Patch, Y. Alter, J. C. Campbell, K. K. Gundersen, J. T. Tang, K. Tsuyuki, J. K. Stockman, The Effect of Intimate Partner Violence and Probable Traumatic Brain Injury on Mental Health Outcomes for Black Women. J Aggress Maltreat Trauma 28, 714–731 (2019).

38. G. Hunnicutt, K. Lundgren, C. Murray, L. Olson, The Intersection of Intimate Partner Violence and Traumatic Brain Injury: A Call for Interdisciplinary Research. J Fam Viol 32, 471–480 (2017).

39. L. A. Fraade-Blanar, B. E. Ebel, E. B. Larson, J. M. Sears, H. J. Thompson, K. C. G. Chan, P. K. Crane, Cognitive Decline and Older Driver Crash Risk. J Am Geriatr Soc 66, 1075–1081 (2018).

40. A. K. Malhotra, R. H. Jaffe, H. Shakil, F. Mathieu, A. B. Nathens, A. V. Kulkarni, C. Diep, E. Y. Yuan, K. S. Ladha, P. C. Coyte, J. R. Wilson, W. P. Wodchis, C. D. Witiw, Unemployment and Personal Income Loss After Traumatic Brain Injury. JAMA Surg 159, 1415–1422 (2024).

41. E. M. Valera, H. Berenbaum, Brain injury in battered women. Journal of Consulting and Clinical Psychology 71, 797–804 (2003).

42. A. E. Tanriverdi, G. L. Iverson, E. M. Valera, Examining Associations Between Intimate Partner Violence-Related Brain Injury, Psychological Abuse, and Cognitive Functioning in Community Women. J Fam Viol, doi: 10.1007/s10896-025-01032-7 (2026).

43. J. E. Karr, S. E. Leong, E. O. Ingram, T. K. Logan, Repetitive Head Injury and Cognitive, Physical, and Emotional Symptoms in Women Survivors of Intimate Partner Violence. Journal of Neurotrauma 41, 486–498 (2024).

44. J. Makovec Knight, O. Hannon, G. Spitz, A. Astridge, C. Copas, B. Duarte Martins, R. Rowse, S. McDonald, C. Padgett, S. Meyer, S. Shultz, G. F. Symons, J. Ponsford, Brain Injury Within Intimate Partner Violence: What Are the Cognitive Effects? Journal of Neurotrauma 43, 269–279 (2026).

45. J. M. Nemeth, C. Decker, R. Ramirez, L. Montgomery, A. Hinton, S. Duhaney, R. Smith, A. Glasser, A. (Abby) Bowman, E. Kulow, A. Wermert, Partner-Inflicted Brain Injury: Intentional, Concurrent, and Repeated Traumatic and Hypoxic Neurologic Insults. Brain Sciences 15, 524 (2025).

46. Y. Zhang, Y. Ma, S. Chen, X. Liu, H. J. Kang, S. Nelson, S. Bell, Long-Term Cognitive Performance of Retired Athletes with Sport-Related Concussion: A Systematic Review and Meta-Analysis. Brain Sciences 9, 199 (2019).

47. H. G. Belanger, R. D. Vanderploeg, The neuropsychological impact of sports-related concussion: A meta-analysis. Journal of the International Neuropsychological Society 11, 345–357 (2005).

48. J. E. Karr, C. N. Areshenkoff, E. C. Duggan, M. A. Garcia-Barrera, Blast-Related Mild Traumatic Brain Injury: A Bayesian Random-Effects Meta-Analysis on the Cognitive Outcomes of Concussion among Military Personnel. Neuropsychol Rev 24, 428–444 (2014).

49. A. D. Witol, F. M. Webbe, Soccer heading frequency predicts neuropsychological deficits. Archives of Clinical Neuropsychology 18, 397–417 (2003).

50. T. McAllister, M. McCrea, Long-Term Cognitive and Neuropsychiatric Consequences of Repetitive Concussion and Head-Impact Exposure. Journal of Athletic Training 52, 309–317 (2017).

51. A. L. C. Schneider, J. R. Pike, H. Elser, J. Coresh, T. H. Mosley, R. Diaz-Arrastia, R. F. Gottesman, Traumatic brain injury and cognitive change over 30 years among community-dwelling older adults. Alzheimer’s & Dementia 20, 6232–6242 (2024).

52. K. M. Guskiewicz, S. W. Marshall, J. Bailes, M. McCrea, R. C. Cantu, C. Randolph, B. D. Jordan, Association between recurrent concussion and late-life cognitive impairment in retired professional football players. Neurosurgery 57, 719–726; discussion 719-726 (2005).

53. National Operating Committee on Standards for Athletic Equipment, Standard Performance Specification for Newly Manufactured Football Helmets (NOCSAE DOC [ND] 002-25, 2025); https://nocsae.org/nd002-standard-performance-specification-for-newly-manufactured-football-helmets/.

54. J. S. Patricios, K. J. Schneider, J. Dvorak, O. H. Ahmed, C. Blauwet, R. C. Cantu, G. A. Davis, R. J. Echemendia, M. Makdissi, M. McNamee, S. Broglio, C. A. Emery, N. Feddermann-Demont, G. W. Fuller, C. C. Giza, K. M. Guskiewicz, B. Hainline, G. L. Iverson, J. S. Kutcher, J. J. Leddy, D. Maddocks, G. Manley, M. McCrea, L. K. Purcell, M. Putukian, H. Sato, M. P. Tuominen, M. Turner, K. O. Yeates, S. A. Herring, W. Meeuwisse, Consensus statement on concussion in sport: the 6th International Conference on Concussion in Sport-Amsterdam, October 2022. Br J Sports Med 57, 695–711 (2023).

55. B. M. Asken, G. D. Rabinovici, Identifying degenerative effects of repetitive head trauma with neuroimaging: a clinically-oriented review. acta neuropathol commun 9, 96 (2021).

56. Q. Boyle, J. Illes, D. Simonetto, P. van Donkelaar, Ethicolegal considerations of screening for brain injury in women who have experienced intimate partner violence. J Law Biosci 9, lsac023 (2022).

57. J. McCleary-Sills, S. Namy, J. Nyoni, D. Rweyemamu, A. Salvatory, E. Steven, Stigma, shame and women’s limited agency in help-seeking for intimate partner violence. Global Public Health 11, 224–235 (2016).

58. M. E. Dichter, K. V. Rhodes, Intimate Partner Violence Survivors’ Unmet Social Service Needs. Journal of Social Service Research 37, 481–489 (2011).

59. C. Lippy, S. N. Jumarali, N. A. Nnawulezi, E. P. Williams, C. Burk, The Impact of Mandatory Reporting Laws on Survivors of Intimate Partner Violence: Intersectionality, Help-Seeking and the Need for Change. J Fam Viol 35, 255–267 (2020).

60. U.S. Census Bureau, Educational Attainment in the United States: 2024 (U.S. Census Bureau, 2025); https://www.census.gov/data/tables/2024/demo/educational-attainment/cps-detailed-tables.html.

61. B. Guerra-Carrillo, K. Katovich, S. A. Bunge, Does higher education hone cognitive functioning and learning efficacy? Findings from a large and diverse sample. PLOS ONE 12, e0182276 (2017).

62. A. Weitzman, Does Increasing Women’s Education Reduce Their Risk of Intimate Partner Violence? Evidence from an Education Policy Reform. Criminology 56, 574–607 (2018).

63. M. Stenberg, A. K. Godbolt, C. Nygren De Boussard, R. Levi, B.-M. Stålnacke, Cognitive Impairment after Severe Traumatic Brain Injury, Clinical Course and Impact on Outcome: A Swedish-Icelandic Study. Behavioural Neurology 2015, 680308 (2015).

64. B. L. Brooks, E. M. S. Sherman, G. L. Iverson, Embedded Validity Indicators on CNS Vital Signs in Youth with Neurological Diagnoses. Arch Clin Neuropsychol 29, 422–431 (2014).

65. R. T. Lange, G. L. Iverson, B. L. Brooks, V. L. Ashton Rennison, Influence of poor effort on self-reported symptoms and neurocognitive test performance following mild traumatic brain injury. Journal of Clinical and Experimental Neuropsychology 32, 961–972 (2010).

66. R. T. Lange, T. A. Brickell, S. M. Lippa, L. M. French, Clinical utility of the Neurobehavioral Symptom Inventory validity scales to screen for symptom exaggeration following traumatic brain injury. Journal of Clinical and Experimental Neuropsychology 37, 853–862 (2015).

67. L. McWhirter, C. W. Ritchie, J. Stone, A. Carson, Performance validity test failure in clinical populations—a systematic review. J Neurol Neurosurg Psychiatry 91, 945–952 (2020).

68. A. S. Ord, R. D. Shura, A. R. Sansone, S. L. Martindale, K. H. Taber, J. A. Rowland, Performance Validity and Symptom Validity Tests: Are They Measuring Different Constructs? Neuropsychology 35, 241–251 (2021).

69. J. J. Roor, M. J. V. Peters, B. Dandachi-FitzGerald, R. W. H. M. Ponds, Performance Validity Test Failure in the Clinical Population: A Systematic Review and Meta-Analysis of Prevalence Rates. Neuropsychol Rev 34, 299–319 (2024).

70. P. H. Montenigro, M. L. Alosco, B. M. Martin, D. H. Daneshvar, J. Mez, C. E. Chaisson, C. J. Nowinski, R. Au, A. C. McKee, R. C. Cantu, M. D. McClean, R. A. Stern, Y. Tripodis, Cumulative Head Impact Exposure Predicts Later-Life Depression, Apathy, Executive Dysfunction, and Cognitive Impairment in Former High School and College Football Players. Journal of Neurotrauma 34, 328–340 (2017).

71. J. Mez, D. H. Daneshvar, B. Abdolmohammadi, A. S. Chua, M. L. Alosco, P. T. Kiernan, L. Evers, L. Marshall, B. M. Martin, J. N. Palmisano, C. J. Nowinski, I. Mahar, J. D. Cherry, V. E. Alvarez, B. Dwyer, B. R. Huber, T. D. Stein, L. E. Goldstein, D. I. Katz, R. C. Cantu, R. Au, N. W. Kowall, R. A. Stern, M. D. McClean, J. Weuve, Y. Tripodis, A. C. McKee, Duration of American Football Play and Chronic Traumatic Encephalopathy. Ann Neurol 87, 116–131 (2020).

72. D. H. Daneshvar, C. J. Nowinski, B. Abdolmohammadi, C. B. Luster, M. Uretsky, B. M. Martin, J. N. Palmisano, J. Weuve, M. D. McClean, J. D. Cherry, B. Dwyer, E. D. Feigel, M. J. Mastrodicasa, V. E. Alvarez, G. D. Rabinovici, W. W. Seeley, L. T. Grinberg, J. F. Crary, T. D. Stein, L. Goldstein, D. I. Katz, R. D. Zafonte, Y. Tripodis, R. C. Cantu, R. A. Stern, M. L. Alosco, A. C. McKee, J. Mez, Prevalence of chronic traumatic encephalopathy at death in National Football League players: retrospective population based cohort study, 2008-21. BMJ 394, e100418 (2026).

73. A. R. Murchland, S. Haneuse, R. B. Lawn, L. Berkman, K. Jakubowski, M. M. Glymour, K. C. Koenen, Intimate partner violence and cognitive functioning – toward quantifying dementia risk. Alzheimer’s & Dementia 21, e70029 (2025).

74. A. M. Kiselica, E. Johnson, J. F. Benge, How impaired is too impaired? Exploring futile neuropsychological test patterns as a function of dementia severity and cognitive screening scores. Journal of Neuropsychology 15, 410–427 (2021).

75. M. T. Bayley, S. Janzen, A. Harnett, R. Teasell, E. Patsakos, S. Marshall, P. Bragge, D. Velikonja, A. Kua, J. Douglas, L. Togher, J. Ponsford, A. McIntyre, INCOG 2.0 Guidelines for Cognitive Rehabilitation Following Traumatic Brain Injury: Methods, Overview, and Principles. J Head Trauma Rehabil 38, 7–23 (2023).

76. J. Ponsford, N. K. Lee, D. Wong, A. McKay, K. Haines, Y. Alway, M. Downing, C. Furtado, M. L. O’Donnell, Efficacy of motivational interviewing and cognitive behavioral therapy for anxiety and depression symptoms following traumatic brain injury. Psychological Medicine 46, 1079–1090 (2016).

77. B. C. Eapen, A. O. Bowles, J. Sall, A. E. Lang, C. W. Hoppes, K. C. Stout, T. Kretzmer, D. X. Cifu, The management and rehabilitation of post-acute mild traumatic brain injury. Brain Injury 36, 693–702 (2022).

78. B. L. Brett, N. Temkin, J. K. Barber, D. O. Okonkwo, M. Stein, Y. G. Bodien, J. Corrigan, R. Diaz-Arrastia, J. T. Giacino, M. A. McCrea, G. T. Manley, L. D. Nelson, for TRACK-TBI Investigators, Long-term Multidomain Patterns of Change After Traumatic Brain Injury: A TRACK-TBI LONG Study. Neurology 101, e740–e753 (2023).

79. N. Glass, K. Laughon, J. Campbell, C. R. Block, G. Hanson, P. W. Sharps, E. Taliaferro, Non-fatal strangulation is an important risk factor for homicide of women. J Emerg Med 35, 329–335 (2008).

80. Violence Against Women Reauthorization Act of 2013, Pub. L. No. 113-4, § 906, 127 Stat. 54 (2013) (codified at 18 U.S.C. § 113(a)(8)).

81. Ohio Senate Bill 288, 134th General Assembly (2022) (codified at Ohio Rev. Code § 2903.18, effective 4 April 2023).

82. Mild Traumatic Brain Injury Committee of the Head Injury Interdisciplinary Special Interest Group, American Congress of Rehabilitation Medicine, Definition of mild traumatic brain injury. J Head Trauma Rehabil 8, 86–87 (1993).

83. M. C. Xu, A. Tanriverdi, G. L. Iverson, E. M. Valera, History of Strangulation Is Associated with Current Traumatic Stress, Self-Reported Vision Problems, and Other Neurobehavioral Symptoms in Women Who Have Experienced Intimate Partner Violence. Journal of Neurotrauma 43, 907–915 (2026).

84. A. R. Macaranas, A. E. Tanriverdi, A.-L. Joseph, G. L. Iverson, E. M. Valera, Pediatric Brain Injuries Are Associated With Intimate Partner Violence-Related Brain Injuries Among Women in Adulthood. J Head Trauma Rehabil 40, 279–286 (2025).

85. A.-L. Joseph Denk, G. L. Iverson, D. P. Terry, E. M. Valera, Mild Brain Injuries Incurred During Intimate Partner Violence Are Related to Objective and Self-Reported Balance Measures. Journal of Neurotrauma 42, 1984–1992 (2025).

86. C. T. Gualtieri, L. G. Johnson, Reliability and validity of a computerized neurocognitive test battery, CNS Vital Signs. Arch Clin Neuropsychol 21, 623–643 (2006).

87. A.-M. G. de Lange, J. H. Cole, Commentary: Correction procedures in brain-age prediction. NeuroImage: Clinical 26, 102229 (2020).

88. C. A. Blevins, F. W. Weathers, M. T. Davis, T. K. Witte, J. L. Domino, The Posttraumatic Stress Disorder Checklist for DSM-5 (PCL-5): Development and Initial Psychometric Evaluation. J Trauma Stress 28, 489–498 (2015).

89. K. Kroenke, R. L. Spitzer, J. B. W. Williams, The PHQ-9. J GEN INTERN MED 16, 606–613 (2001).

90. R. L. Spitzer, K. Kroenke, J. B. W. Williams, B. Löwe, A Brief Measure for Assessing Generalized Anxiety Disorder: The GAD-7. Arch Intern Med 166, 1092–1097 (2006).

91. J. B. Saunders, O. G. Aasland, T. F. Babor, J. R. de la Fuente, M. Grant, Development of the Alcohol Use Disorders Identification Test (AUDIT): WHO Collaborative Project on Early Detection of Persons with Harmful Alcohol Consumption--II. Addiction 88, 791–804 (1993).

92. D. P. Bernstein, J. A. Stein, M. D. Newcomb, E. Walker, D. Pogge, T. Ahluvalia, J. Stokes, L. Handelsman, M. Medrano, D. Desmond, W. Zule, Development and validation of a brief screening version of the Childhood Trauma Questionnaire. Child Abuse Negl 27, 169–190 (2003).

93. K. Hegarty, M. Sheehan, C. Schonfeld, A Multidimensional Definition of Partner Abuse: Development and Preliminary Validation of the Composite Abuse Scale. Journal of Family Violence 14, 399–415 (1999).

94. M. Ford-Gilboe, C. N. Wathen, C. Varcoe, H. L. MacMillan, K. Scott-Storey, T. Mantler, K. Hegarty, N. Perrin, Development of a brief measure of intimate partner violence experiences: the Composite Abuse Scale (Revised)-Short Form (CASR-SF). BMJ Open 6, e012824 (2016).

95. E. von Elm, D. G. Altman, M. Egger, S. J. Pocock, P. C. Gøtzsche, J. P. Vandenbroucke, STROBE Initiative, The Strengthening the Reporting of Observational Studies in Epidemiology (STROBE) statement: guidelines for reporting observational studies. J Clin Epidemiol 61, 344–349 (2008).

96. R. L. Wasserstein, A. L. Schirm, N. A. Lazar, Moving to a World Beyond “p < 0.05.” The American Statistician 73, 1–19 (2019).

